# Assessing sustainable and healthy diets in large-scale surveys: validity and applicability of a dietary index based on a brief food group propensity questionnaire representing the EAT-Lancet planetary health diet

**DOI:** 10.1101/2025.02.07.25321862

**Authors:** Agustin Miranda, Anna Maria Murante, Federica Manca, Fabio Consalez, Anant Jani, Fabrice DeClerck, Matthieu Maillot, Eric Verger

**Author notes:** **Corresponding author:** Agustin MIRANDA, French National Research Institute for Sustainable Development, France.

## Abstract

**Introduction:** Ensuring healthy diets within planetary boundaries is essential. However, current instruments measuring adherence to the EAT-Lancet planetary health diet are unsuitable for large-scale surveys. Simplified tools assessing consumption frequency can improve response rates, lower costs, and facilitate administration. This study aimed to develop a practical and concise index for evaluating relative adherence to the EAT-Lancet diet across large-scale multi-country surveys.

**Methods:** First, the EAT-Lancet Consumption Frequency Index (ELFI) was developed using a brief food propensity questionnaire of 14 food groups representing the planetary health diet from the FEAST survey, which encompassed 27 European countries (n□=□27,417). Subsequently, ELFI was further validated using 24-hour dietary recalls from the INCA3 survey (n = 1,645), correlating it with the valid EAT-Lancet Index (ELI), which evaluates absolute adherence, as well as with food group consumption, measures of nutritional health (nutrient adequacy and diet quality), and environmental impact. Analyses included assessment of reliability, structural validity, concurrent validity, and nomological validity.

**Results:** ELFI showed strong reliability (alpha > 0.80) and factor analysis revealed a two-factor solution: “foods to encourage” and “foods to balance and to limit”. Confirmatory factor analysis demonstrated that ELFI is structurally valid. Concurrent validity was confirmed as it was associated with sex, age, education, income, household size, physical activity and smoking habit (p < 0.05). ELFI correlated with ELI (0.44, p < 0.0001) and food group consumptions. Regarding nomological validity, the ELFI subscores for “foods to encourage” and “foods to balance and to limit” were associated with better nutritional health (β = 0.62 and 0.23, respectively; p < 0.0001) and a lower environmental impact (β = −0.16 and −0.36, respectively; p < 0.0001).

**Conclusion:** ELFI approach represents a valuable and easy-to-implement index for evaluating relative adherence to sustainable and healthy diets in large-scale multi-country studies.

## 1. Introduction

Contemporary food systems have substantial impacts on both human and environmental health (1). As a result, transitioning toward healthier and more sustainable diets is essential for addressing the health and environmental challenges associated with current food production methods and consumption patterns (2,3). In 2019, the EAT-Lancet Commission introduced the planetary health diet, which specifies ranges for food groups to support healthy diets within planetary resource boundaries (4). This diet encourages the consumption of vegetables, fruits, legumes, whole grains, nuts, and fish, while limiting red meats, tubers, saturated fats, and added sugars. Moderate consumption of eggs, poultry, and dairy foods is also advised (4). Despite criticisms regarding cultural acceptance, the risk of nutritional inadequacy, unaffordability for low-resource settings, and its individual-centered approach rather than addressing systemic issues, the planetary health diet remains an important strategy for sustainability, offering a valuable roadmap for transitioning to diets that benefit both human health and the environment (5).

Since the publication of the EAT-Lancet planetary health diet guidelines, there has been a growing research interest in assessing population compliance with these recommendations (6). In this contest, consumers have a pivotal role as they have own food-consumption and purchasing preferences and are targeted by constant stimuli from industry as well. Understanding what, how, and why people eat is essential for enhancing individuals’ food literacy, it makes possible finding new strategies to meet food needs in a healthy and sustainable way, while collectively promoting social change, reducing food waste, and transforming food systems to benefit both human health and climate goals (7). Examining adherence to planetary health diet guidelines also helps to identify the factors that enable or hinder the adoption of sustainable dietary practices. Consequently, several EAT-Lancet-based dietary indices have been developed, applying traditional nutritional assessment methods such as dietary recall, food frequency questionnaires (FFQs), and food diaries to quantify dietary intake and score adherence based on recommended cut-offs (7, 8).

Studies on the validity of EAT-Lancet-based dietary indices and their relationships to health and environmental outcomes have emerged (9, 10–12). However, existing methods may prove impractical for large-scale surveys. First, traditional methods for quantifying food intake are technically complex and challenging to collect, process, and analyze (13, 14). Furthermore, from a financial perspective, these methods require substantial investments to cover the costs of data collection, processing, and analysis (13, 15). This situation becomes even more complicated when applied to multi-country studies involving large populations, as the associated time and cost issues are amplified (16). Additionally, from the respondents’ perspective, the use of these techniques can lead to participant burden, especially in multidimensional surveys designed to address a wide range of topics (17). Therefore, it is essential to develop more cost-effective and simpler ways to implement methods for such studies. Food propensity questionnaires (FPQs), also known as non-quantitative FFQs, represent a valuable alternative that can reduce costs, simplify administration, and improve response rates (18). FPQs are designed to evaluate food consumption patterns and preferences by collecting information on the frequency of consumption for specific food groups (e.g., fruits) or items (e.g., oranges), thereby facilitating the analysis of dietary habits and the factors influencing food choices without requiring precise quantitative measures (19). FPQs are often tailored into brief formats to assess specific dietary components and have been validated against quantitative dietary assessment methods, making the FPQ approach particularly useful in large-scale studies (20). Moreover, data derived from FPQs are useful for developing valid dietary indices (21–24).

Given the limitations of current indices for large-scale studies, the need for streamlined assessment tools is evident (25, 26). Dietary indices should reliably capture adherence to dietary guidelines and demonstrate validity through consistent correlations with relevant health and environmental outcomes (27). A novel metric should correlate with previous validated indices for assessing absolute adherence to the planetary health diet, such as the EAT-Lancet dietary index (ELI). The ELI, developed using FFQ and food diary data from Sweden (28), provides a solid basis for designing FPQ-based indices as it was created with quantitative FFQ data, incorporates 14 food groups categorized into emphasized and restricted categories representing the planetary health diet, uses a simple graded scoring system for each food group (i.e., from 0 to 3 points) which is easy to replicate for large-scale surveys, and it is associated with improved health outcomes and reduced environmental impact (6). This study aimed to develop and validate the EAT-Lancet Consumption Frequency Index (ELFI), a brief and practical index based on FPQs to assess relative adherence to the planetary health diet in large-scale multi-country surveys.

## 2. Methods

### 2.1. Study design and participants

The study included two samples. For the first sample, data were obtained from the FEAST (Food systems that support transitions to hEalthy And Sustainable dieTs) project, which involved a large-scale multi-country sample (29). The FEAST project employed a quantitative cross-sectional design to collect primary data from 27,000 adults (aged 18 years and older) across 27 European countries. A stratified sampling design guided participant enrollment, targeting at least 1,000 individuals, representative of age, gender, and education in each selected country. To achieve this sample, FEAST utilized a market-insights platform, ensuring representativity through demographic quotas and respondents were recruited among national panelists. Each participant voluntarily completed a multidimensional web survey consisting of 88 items, organized into eight sections covering the following topics: dietary patterns; purchasing and consumption behaviors; perceptions of sustainability and sources of information; drivers and barriers to healthy and sustainable diets; perceptions on various food policies; and sociodemographic information. Ethical approval for the FEAST survey was granted by the conjoined Ethics Committee of Scuola Superiore Sant’Anna and Scuola Normale Superiore (Resolution No. 29/2023, and following amendments). A detailed description of the methodology and study design of the FEAST survey can be found elsewhere (30).

For the second phase of instrument validation, data were extracted from the French Third Individual and National Study on Food Consumption Survey (INCA3), which provides both 24-hour dietary recall and FPQ useful for the index validity assessment (31). This nationally representative cross-sectional study involved 4,114 individuals in mainland France from February 2014 to September 2015. A detailed description of the methodology and study design of INCA3 can be found elsewhere (31). For the current study, participants aged 18 years and older were included in the analysis. Individuals who provided both 24-hour recall and FFQ data were considered eligible. Mis-reporters were excluded based on discrepancies between their reported energy intake and estimated basal metabolic rate. Consequently, the final sample consisted of 1,645 adults, comprising 690 men and 955 women. The INCA3 study adhered to the Declaration of Helsinki and received approval from the French Data Protection Authority (Decision DR 2013-228) and the Advisory Committee on Information Processing in Health Research (Opinion 13.055). Verbal informed consent was obtained from all participants.

### 2.2. Index development and validation

#### 2.2.1. Measuring the relative adherence to the planetary health diet in a large-scale survey

To assess dietary behaviors across Europe, the FEAST survey developed a brief FPQ representing the EAT-Lancet planetary health diet (30). This questionnaire evaluated the consumption of 14 key food groups outlined in the planetary health diet, including whole grains, tubers and starches, vegetables, fruits, dairy foods, red meats, poultry, eggs, fish and shellfish, legumes, unsaturated oils, animal fats and saturated oils, and added sugars (4). Participants reported their consumption of these food groups using seven frequency categories: never, once-to-three times per month, once per week, two-to-three times per week, four-to-six times per week, daily (once a day), and two or more times daily.

A quantile-based scoring system was subsequently developed. This approach enables relative scoring based on the distribution of consumption observed within the study population, such as quartiles or tertiles (32). The food groups covered in the FEAST survey, along with their specifications about food items and the complete scoring system, are shown in Figure 1. Whole grains, vegetables, fruits, fish and shellfish, legumes, nuts, and plant oils were classified as foods to encourage, while tubers, dairy foods, red meats, eggs, and poultry were categorized as foods to balance. In contrast, animal fats and added sugars were designated as foods to limit. This classification aligns with the criteria of the EAT-Lancet framework; however, in this study, red meats and tubers were considered as foods to balance rather than foods to limit. Accordingly, other EAT-Lancet-based dietary indices have adopted these modifications in nutrient-rich food groups to account for micronutrient adequacy (6, 33).

**Figure 1.**
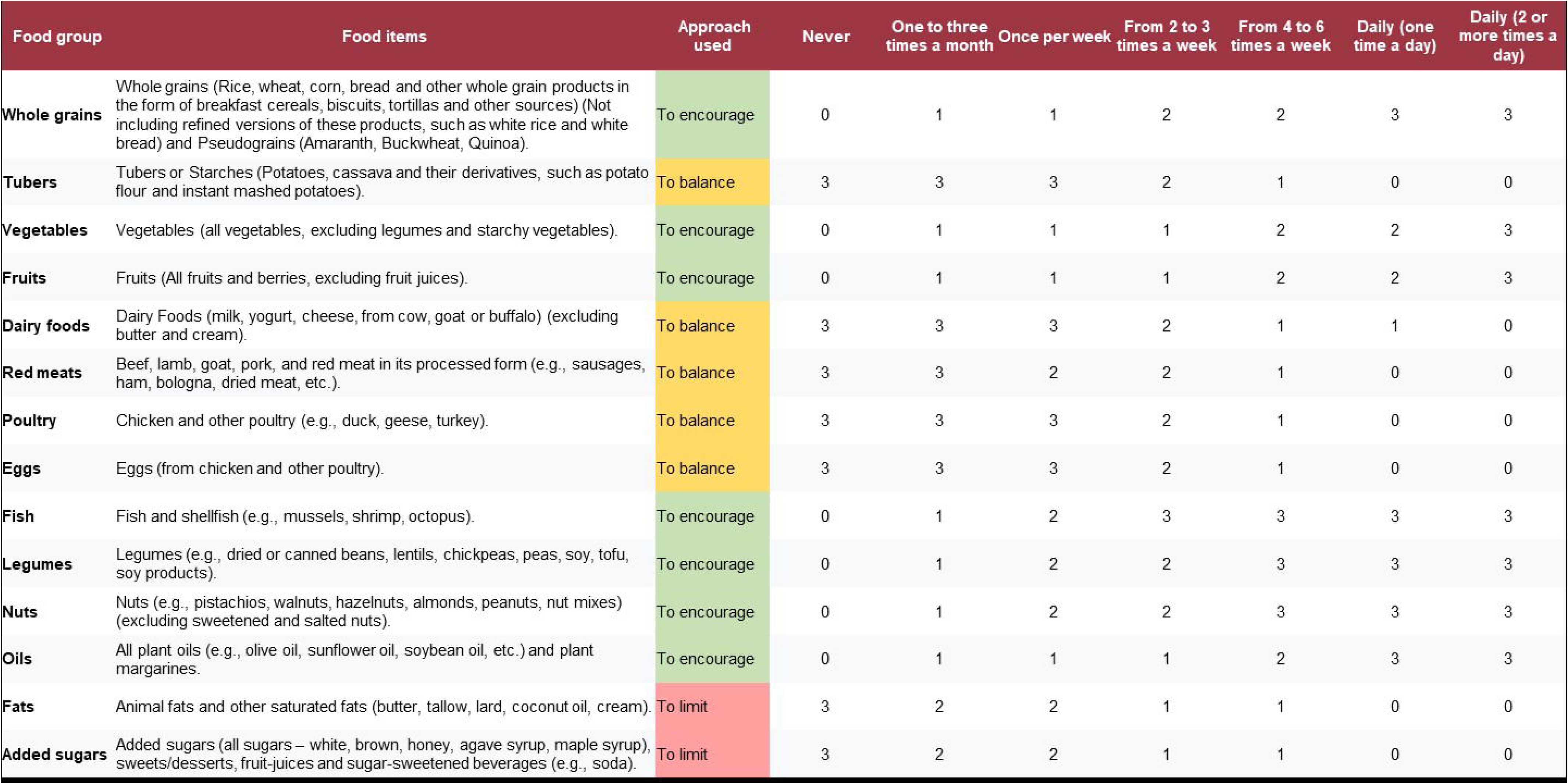
Scoring system used in the EAT-Lancet Consumption Frequency Index (ELFI) for assessing relative adherence to the Planetary Health Diet in the FEAST large-scale multi-country survey.

Figure 2 provides a detailed description of the scoring procedure. Briefly, food groups to encourage were scored according to tertiles, with points ranging from 1 to 3 based on consumption frequency. Specifically, 1 point was assigned to the first tertile (indicating low consumption frequency) and 3 points to the third tertile (indicating high consumption frequency). Individuals who reported not consuming these food groups received 0 points. In contrast, for foods to limit and to balance, the scoring system was inverted, meaning a higher score indicated lower consumption frequency. Furthermore, for tubers, dairy foods, red meats, poultry, and eggs, a nuanced and flexible scoring approach was employed, with cut-offs based on interdisciplinary expert consensus. This method considered minimum consumption levels while avoiding the imposition of absolute restrictions on nutrient-rich foods, also ensuring that adherence does not disproportionately favor specific dietary patterns, such as veganism or vegetarianism, over other balanced diets. The use of minimum intake levels is a common approach in the development of EAT-Lancet based indices to ensure that adherence supports adequate nutrient intake, which is one of the main challenges of the planetary health diet (6, 33). For tubers, chicken, and eggs, a frequency of never to once a week received 3 points, two to three times per week received 2 points, four to six times per week received 1 point, and consumption once a day or more received 0 points. In the case of dairy foods, participants received 3 points if they consumed them never to once a week, 2 points for two to three times per week, 1 point for four times per week to once a day, and 0 points for more than once a day. Red meats group was scored as follows: never to three times per month received 3 points, one to three times per week received 2 points, four to six times per week received 1 point, and once a day or more received 0 points. The total ELFI score was calculated as the sum of its 14 food component scores, with a theoretical range of 0 to 42. A higher score indicates higher relative adherence to the EAT-Lancet planetary health diet.

**Figure 2.**
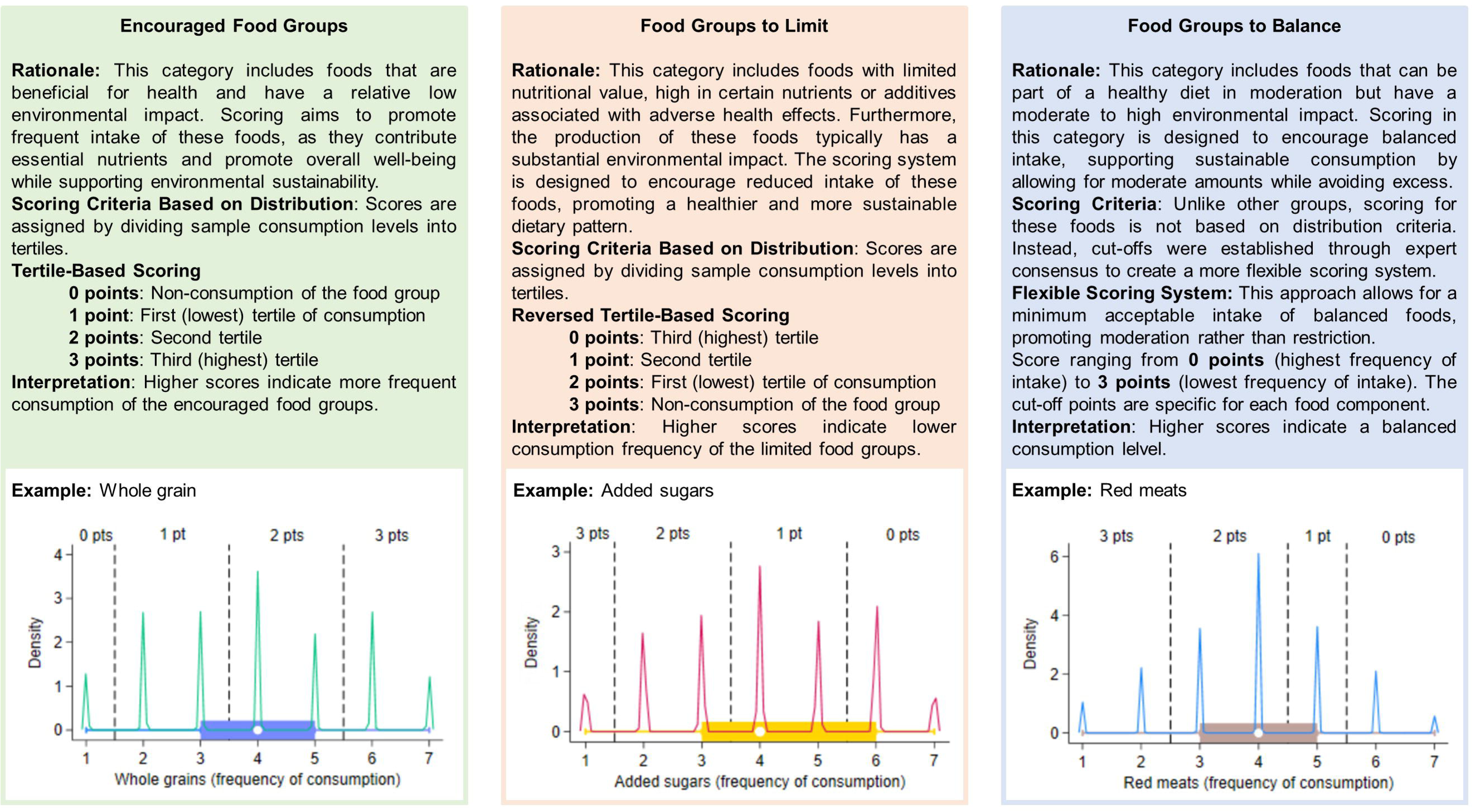
Scoring procedure for the EAT-Lancet Consumption Frequency Index (ELFI). The graphs display the distribution of consumption frequency responses, represented as density curves and box plots, with a scale ranging from 1 (never) to 7 (two or more times a day). Density represents how concentrated data points are within different ranges of the consumption, indicating the distribution of the data across the frequencies.

#### 2.2.2. Calculating ELFI in INCA3

The same approach and tertile-based scoring system in the FEAST study were replicated in INCA3 survey to assess index validity. INCA3 survey employed a FPQ featuring 75 food items selected based on three key criteria: their contribution to total nutrient and chemical intake, their impact on inter-individual variability in total intake, and a significant increase in consumer rates observed in the three-day food record compared to the seven-day record from the previous INCA2 survey (31, 34). Participants reported their consumption status for each food item with a yes/no response and indicated their frequency of consumption over the past twelve months. To standardize food intake frequency, the reported number of days per month were transformed into seven categorical frequency scales from “never” to “every day”. The complete list of foods included in the FPQ used in INCA3 is available at https://www.data.gouv.fr/fr/datasets/r/bebbad5e-4fad-46ca-ab5e-6edc9966c374.

#### 2.2.3. Assessing the validity of the ELFI against 24-hour recall

Data derived from the 24-hour recalls of the INCA3 survey were used to calculate the EAT-Lancet Index (ELI), a validated measure based on consumption quantity to assess absolute adherence to the planetary health diet that serves as a benchmark for testing ELFI validity (28). Dietary intake was assessed using three non-consecutive 24-hour dietary recalls, which included two weekdays and one weekend day over a three-week period (31). Participants were contacted by phone to report all foods and beverages consumed during the previous day, utilizing validated photographs of standard portion sizes in France. The energy and nutrient content of the foods were derived from the 2016 database of the French Centre d’Information sur la Qualité des Aliments (35). Traditional recipes and dishes containing various ingredients were disaggregated into their components based on average recipes obtained from existing recipe database and popular cooking sources in France, such as marmiton.org. In this study, the dietary data were used to calculate adherence indices for the EAT-Lancet diet, along with nutritional quality and environmental scores for each individual participating in the INCA3 survey.

The ELI comprises 14 food groups divided into two categories: seven positive groups, referred to as “emphasized foods” (vegetables, fruits, unsaturated oils, legumes, nuts, whole grains, and fish), and seven negative groups, known as “limited foods” (beef and lamb, pork, poultry, eggs, dairy, potatoes, and added sugar) (28). The alignment of dietary intake, measured in grams per day (g/d, without adjustments for daily energy), with EAT-Lancet recommendations is evaluated using a scoring system based on a graded scale that ranges from 0 (indicating non-compliance) to 3 points (indicating high compliance). Total scores for the ELI dietary index can range from 0 to 42 points. Further details regarding the ELI index, including scoring criteria and cut-off points, are provided in Supplementary Materials and elsewhere (28).

#### 2.2.4. Assessing nutritional health and environmental indicators

Since the planetary health diet has been designed to be both healthy and sustainable, it is crucial to evaluate whether the index adequately correlates with nutritional health and environmental impact indicators. Different dimensions of nutritional health were assessed, including nutrient adequacy, diet quality, and the diet’s inflammatory and antioxidant profile. In this sense, nutrient adequacy ensures the diet meets essential physiological needs, preventing deficiencies or excesses that could harm health (36). The diet quality refers to overall balance of the diet, promoting nutrient-dense, diverse foods while minimizing unhealthy components, ensuring the index supports a well-rounded and healthful dietary pattern (37,38). The inflammatory and antioxidant profile further assesses the diet’s role in reducing inflammation and oxidative stress, which are linked to chronic disease risk, ensuring that the index reflects dietary patterns that support both short and long-term health outcomes (39–41).

Nutrient adequacy was assessed using the PANDiet score (36). The PANDiet assesses the likelihood of meeting recommended intake levels for 33 nutrients through two sub-scores: adequacy and moderation. The adequacy sub-score averages probabilities for 27 nutrients, including proteins, carbohydrates, dietary fiber, essential fatty acids, vitamins, and minerals, while the moderation sub-score focuses on limiting intake of 6 nutrients, such as sugars, saturated fats, and sodium. Each sub-score is multiplied by 100, and their average yields the total PANDiet score, ranging from 0 to 100. Higher scores indicate better diet quality and greater nutrient adequacy, based on dietary reference values from the French Agency for Food, Environmental and Occupational Health and Safety (ANSES). Additional information about PANDiet is available in Supplementary Materials.

Additionally, the Global Diet Quality Score (GDQS) was used for assessing diet quality (37). The GDQS is based on 25 food categories important for nutrient supply and reducing chronic disease risk. It consists of 16 healthy food components (higher consumption increases the score), 7 unhealthy food components (higher consumption decreases the score), and 2 components that are unhealthy in excess (resulting in a score of 0 for both insufficient and excessive intake). The total GDQS score ranges from 0 to 49 points and includes two sub-scores: GDQS+, which reflects healthy food components (0 to 32 points), and GDQS-, which reflects unhealthy or over-consumed food components (0 to 17 points). In addition, the comprehensive Diet Quality Index (cDQI) was used to evaluate diet quality by distinguishing between plant-based and animal-based food components that are considered either beneficial or detrimental to health (38). The cDQI consists of 11 plant-based components (pDQI) and 6 animal-based components (aDQI). Healthy foods receive positive scores, while unhealthy foods are given negative scores. The total cDQI score ranges from 0 to 85. Additional information about GDQS and cDQI is available in Supplementary Materials.

The Dietary Inflammatory Index (DII) was used to evaluate the inflammatory potential of the diet (39). For this study, 34 dietary parameters (including foods and nutrients) associated with inflammatory biomarkers were used to calculate the DII score. The DII calculation involves the following steps: dietary intake is compared to a global reference standard, Z-scores are computed for each dietary component, and these scores are converted into centered proportions. The centered values are then multiplied by inflammatory effect scores specific to each parameter, and the results are summed to generate the total DII score. A higher DII score reflects a more proinflammatory diet. Detailed information on the specific steps involved in the calculation can be found elsewhere and in Supplementary Material (40).

The Composite Dietary Antioxidant Index (CDAI) was calculated to evaluate overall exposure to dietary antioxidants (41). This index incorporates key antioxidants such as vitamins A, C, and E, as well as manganese, selenium, and zinc, providing a measure of an individual’s dietary antioxidant profile. To calculate the CDAI, each antioxidant was standardized by subtracting the sex-specific mean and dividing by the sex-specific standard deviation, and then the standardized values were summed. A higher CDAI score indicates greater antioxidant availability in the diet, suggesting enhanced potential for defense against oxidative stress and improved health protection (42). More information is available in Supplementary Materials.

The environmental impact analysis was conducted using the Agribalyse 3.1.1 database, developed by the French Agency for the Environment and Energy Management (43). Agribalyse 3.1.1 provides reference data on the environmental effects of agricultural and food products, employing the Life Cycle Assessment (LCA) methodology, which accounts for various stages of the food supply chain. In this study, key environmental indicators were used, including greenhouse gas emissions (kg CO2 eq), exposure ionizing radiation (kg U235 eq), photochemical ozone formation (kg NMVOC eq), ozone depletion (Freon-11), emission of particulate matter in change (mortality due to particulate matter emissions), acidification (mol H+ eq), terrestrial eutrophication (mol N eq), freshwater eutrophication (kg P eq), marine eutrophication (kg N eq), freshwater ecotoxicity in (CTUe), water use (m3 world eq), land use (loss of soil organic matter content in kg carbon deficit), fossils resource use (MJ), metals and minerals resource use (kg Sb eq), and the aggregated score product environmental footprint. The complete methodology for the Agribalyse 3.1.1 database is described elsewhere (44).

The sociodemographic variables analyzed included sex (woman or man), age, and educational level (categorized as no diploma or primary school diploma, lower secondary school diploma, high-school to 2 years of higher education, 3 or more years of higher education) (31). Respondents provide information about household size (number of persons) and income per consumption unit (CU), which was classified as less than €900/month, €900-€1340/month, €1340-€1850/month, and €1850 or more per month. Body Mass Index (BMI) was calculated by dividing the individual’s self-reported weight in kilograms by the square of their height in meters and classified according to WHO BMI categories (i.e., underweight, normal, overweight, and obesity) (45). A modified version of the Recent Physical Activity Questionnaire assessed adults’ physical activity and sedentary behavior over the past four weeks, covering leisure, household, and work activities (31, 46). Analysis included activity frequency, duration, and intensity (in metabolic equivalent of task-METs), producing indicators such as weekly energy expenditure in MET/minutes, daily sedentary hours, and activity levels relative to WHO recommendations. Based on these metrics, individuals were classified as: inactive and sedentary (activity below recommendations and sedentary >7 h/d), inactive and non-sedentary (activity below recommendations and sedentary ≤7 h/d), active and sedentary (activity meets recommendations and sedentary >7 h/d), or active and non-sedentary (activity meets recommendations and sedentary ≤7 h/d) (47).

### 2.3. Statistical analysis

Statistical analyses were performed using Stata (version 18, StataCorp, College Station, TX, USA) and the threshold for statistical significance was p < 0.05. Mean and standard deviation (SD) were calculated for numerical variables, and percentages for categorical ones. The INCA3 survey weighting factors were used. The evaluation of the index’s metric properties included the examination of convergent validity, concurrent-criterion validity, nomological validity, and structural validity, following the well-established approaches for dietary index development and validation (25–27).

#### 2.3.1. Structural validity

Structural validity assesses whether the food groups of an index are appropriately organized and effectively capture the underlying dimensions intended for evaluation (25). To assess reliability and structural validity, the study used Cronbach’s alpha, as well as exploratory factor analysis (EFA) and confirmatory factor analysis (CFA) in the FEAST survey sample (n = 27,417). Cronbach’s α measures the internal consistency reliability, reflecting how well the food groups within the index correlate with each other (25). A value of α greater than 0.70 indicates that the food groups work together effectively to reliably measure the same underlying dimension.

Subsequently, the survey sample was randomly divided into two halves, with one subsample used for EFA (n = 13,709) and the other one for CFA (n = 13,708). Briefly, the purpose of EFA was to identify an optimal factor structure, while CFA aimed to confirm the dimensionality of this structure (48). EFA was performed using varimax-rotated principal component factor analysis. The Kaiser-Meyer-Olkin (KMO) test and Bartlett’s test of sphericity were used to assess the data adequacy. Factors were retained based on an eigenvalue greater than 1.0 and the identification of the elbow point in the scree plot, which marks the point where the eigenvalues begin to flatten, indicating the number of factors that explain the most variance in the data. A factor loading threshold of 0.30 was used to determine meaningful contributions to the latent factor.

CFA was conducted on the second half-split subsample to validate the factor structure identified in EFA. Maximum-likelihood structural equation modeling (SEM) were used in the CFA, and goodness-of-fit indices were calculated for both unidimensional and bidimensional models. These indices included the Root Mean Square Error of Approximation (RMSEA), Comparative Fit Index (CFI), Standardized Root Mean Square Residual (SRMR), AIC (Akaike Information Criterion), BIC (Bayesian Information Criterion), and Coefficient of Determination (CD). Modification indices were used to guide model improvement. Finally, a bifactor model was tested to determine whether the index was sufficiently unidimensional to justify using a total score, while still accounting for the multidimensionality identified during the analysis.

#### 2.3.2. Convergent validity

In dietary index development, convergent validity refers to the extent to which a dietary index correlates with other established measures that assess the same or similar dietary construct (27). Convergent validity is demonstrated through correlations between the new index and pre-existing valid metrics, testing the accuracy and reliability of the index as a measure of the targeted dietary constructs. This study calculated the correlation between the ELFI (based on FPQ responses of INCA3) and ELI (based on 24h-recalls of INCA3), employing Pearson’s r coefficient for total scores and Spearman’s ρ coefficients for each equivalent food groups. Additionally, ρ between the ELFI and the quantity of food consumed was also analyzed. The ability of ELFI to assess diet quality independently of diet quantity, measured by dietary energy intake, was also evaluated. For this purpose, a regression analysis was conducted to examine the relationship between total daily kilocalories (excluding alcohol) and ELFI, adjusted for sex and age. Participants were ranked and categorized into quintiles (Q1 to Q5) of ELFI, with Q1 representing the lowest level of relative adherence and Q5 representing the highest. Food consumption (g/d) was compared across quintiles using ANOVA, and trends were identified through the Jonckheere-Terpstra test.

#### 2.3.3. Concurrent-criterion validity

Concurrent-criterion validity assesses how well a dietary index correlates with external criteria and its ability to differentiate between groups known to exhibit differences in diet quality (49). Thus, this metric property is analyzed by comparing index scores across various sociodemographic groups that are expected to have distinct dietary patterns. In this study, concurrent-criterion validity was evaluated using multiple linear regression, with ELFI score as the dependent variable and sociodemographics as predictor variables. The results were expressed as coefficient plots, including 95% confidence intervals and adjusted β values.

#### 2.3.4. Nomological validity

Nomological validity refers to the degree to which a construct fits within a broader theoretical framework and exhibits expected relationships with other related constructs (50). It involves assessing whether the dietary index behaves as predicted in relation to other established variables based on theory. Thus, as ELFI intends to measure a healthy and sustainable diet, nomological validity involved examining how it correlates with nutritional health outcomes and environmental impact indicators. SEM was used to test nomological validity, which combines elements of multiple regression and factor analysis to explore the structural relationships between measured variables and latent variables (constructs) (51). The nomological model comprised four latent variables: “foods to encourage” (including whole grains, vegetables, fruits, fish and shellfish, and legumes), “foods to balance or limit” (including tubers, dairy foods, red meats, poultry, eggs, and added sugars), “nutritional health” (represented by PANDiet, GDQS, cDQI, DII, and CDAI), and “environmental impact” (represented by greenhouse gas emissions, particulate matter emissions, terrestrial eutrophication, water use, and land use). The five environmental indicators were selected based on their relevance, as maintaining an equal number of measured variables for each latent outcome variable enhances model parsimony, identification, and interpretability. The model was estimated using maximum likelihood estimation with the Satorra-Bentler correction, and standardized effects were expressed as β coefficients. Goodness-of-fit indices were calculated to assess model fit the data.

Additionally, Spearman’s bivariate correlations were analyzed between ELFI and the probability of nutritional adequacy for all nutrients measured through PANDiet, as well as with all indicators of nutritional quality and environmental impact, with results presented in radar plots.

## 3. Results

The average ELFI score within the FEAST sample was 23.71 (3.60). Among the food groups, poultry and tubers had the highest mean scores of 2.04 (0.99) and 2.29 (1.01), respectively, whereas added sugars and fats had the lowest mean scores, with values of 1.19 (0.90) and 1.30 (0.88), respectively. Reliability analysis showed that the item-total correlations for all food groups ranged from 0.44 to 0.65, while the item-rest correlations ranged from 0.33 to 0.55 (Table 1). The overall ELFI yielded an α of 0.83, indicating strong internal consistency reliability. Similarly, “Foods to encourage” and “Foods to balance and to limit” subscores exhibited adequate reliability levels (α = 0.74 and α = 0.75, respectively). Furthermore, no improvement in α was observed upon the removal of any individual food group, further supporting the reliability of the index. These findings suggest that the food groups within ELFI exhibit strong internal consistency and homogeneity, while avoiding redundancy.

**Table 1.**
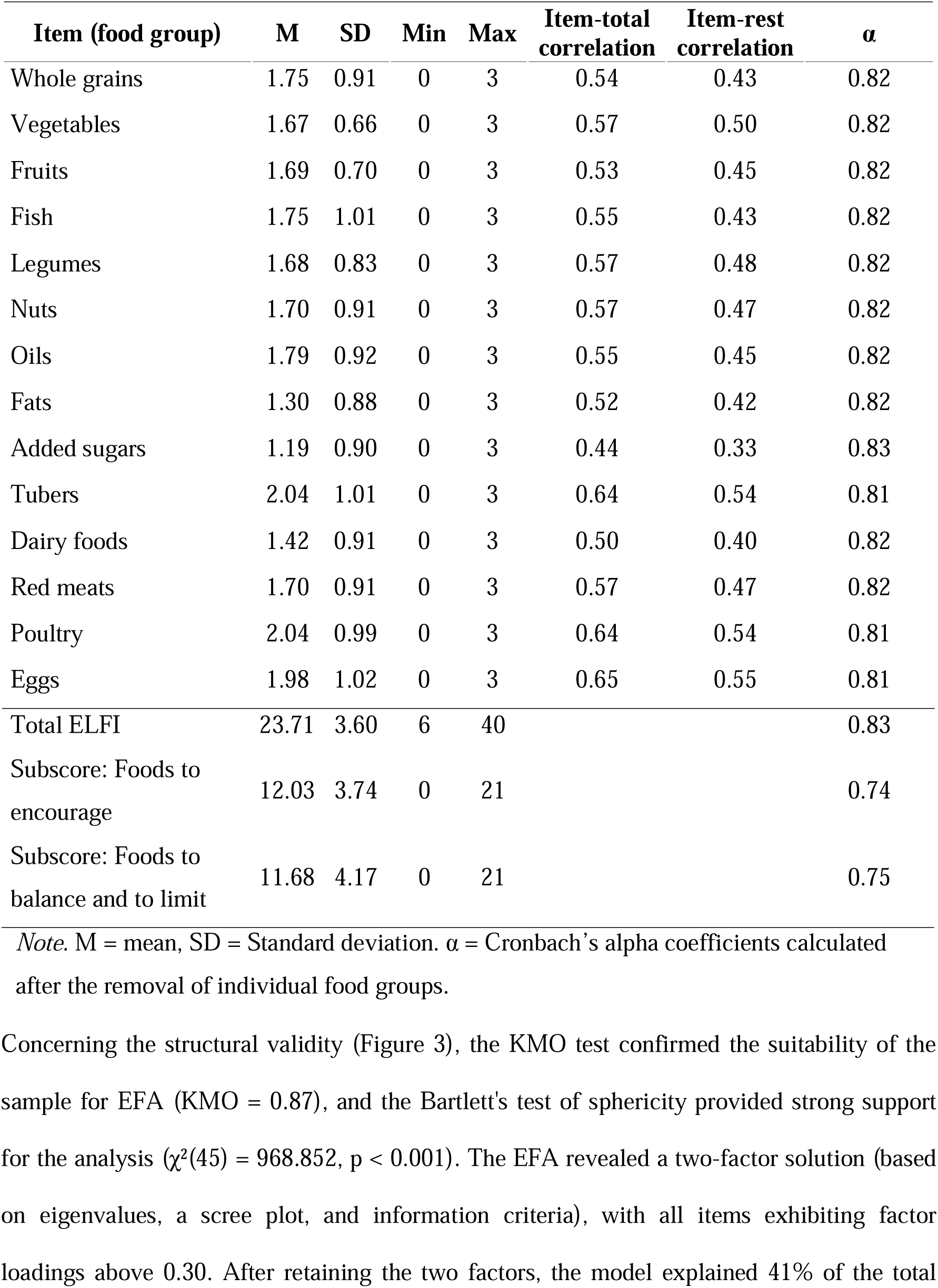
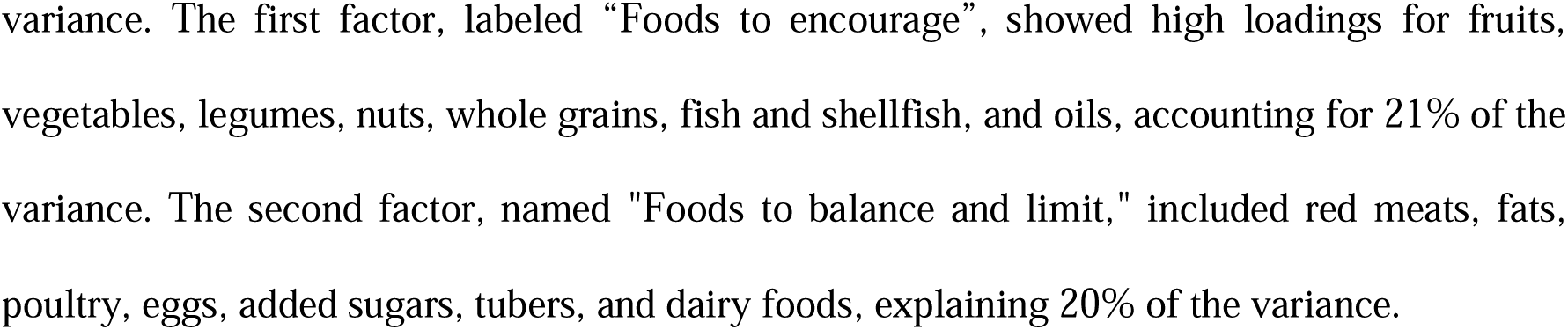
Reliability analysis of the EAT-Lancet Consumption Frequency Index (ELFI) in the FEAST survey (n = 27,417).

Concerning the structural validity (Figure 3), the KMO test confirmed the suitability of the sample for EFA (KMO = 0.87), and the Bartlett’s test of sphericity provided strong support for the analysis (χ²(45) = 968.852, p < 0.001). The EFA revealed a two-factor solution (based on eigenvalues, a scree plot, and information criteria), with all items exhibiting factor loadings above 0.30. After retaining the two factors, the model explained 41% of the total variance. The first factor, labeled “Foods to encourage”, showed high loadings for fruits, vegetables, legumes, nuts, whole grains, fish and shellfish, and oils, accounting for 21% of the variance. The second factor, named “Foods to balance and limit,” included red meats, fats, poultry, eggs, added sugars, tubers, and dairy foods, explaining 20% of the variance.

**Figure 3.**
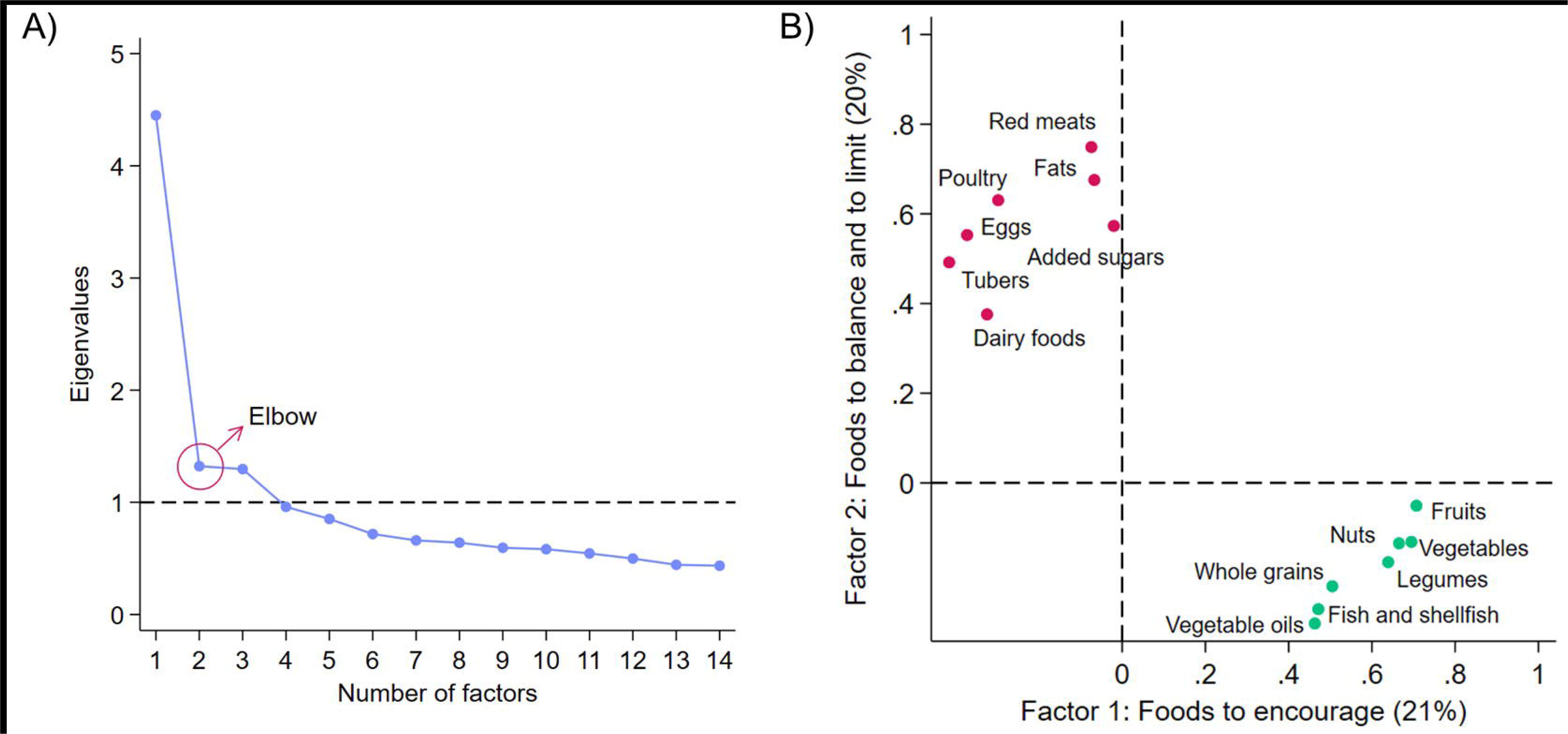
Scree plot showing eigenvalues (A) and rotated factor loading plot (B) obtained from the Exploratory Factor Analysis of the EAT-Lancet Consumption Frequency Index (ELFI) in the FEAST survey (n = 27,417). Percentages in parentheses indicate the variance explained after retaining two factors.

Table 2 compares the goodness-of-fit measures from the CFA for both unidimensional and bidimensional ELFI models. The results confirmed that ELFI has a bidimensional structure, though both models demonstrated an acceptable data fit. All fit indices met the recommended values based on standard cut-off criteria. Additionally, the CFA supported a bifactor model.

**Table 2.**
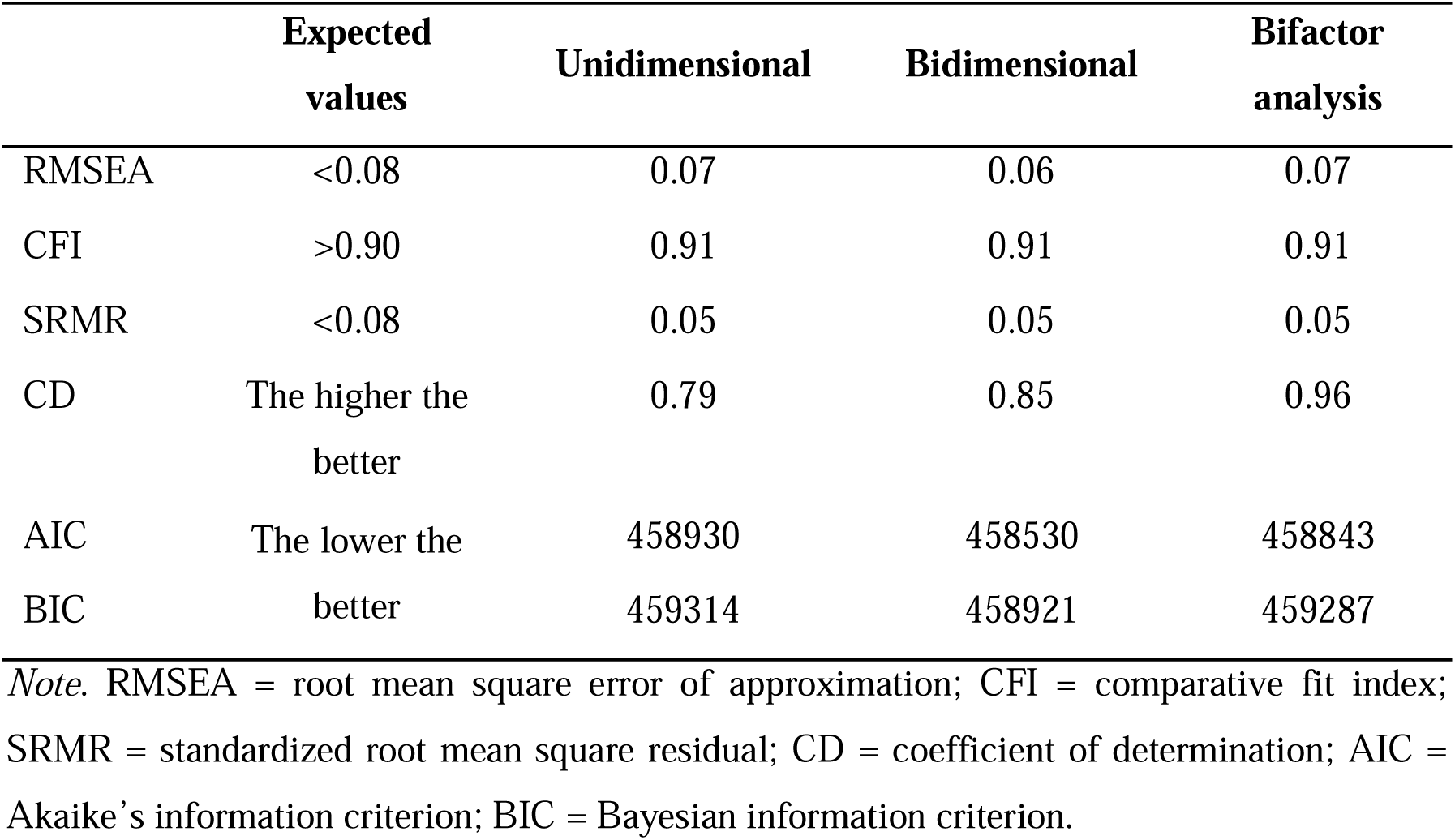
Fit indices for the confirmatory factor analysis models of the EAT-Lancet Consumption Frequency Index (ELFI) in the FEAST survey (n = 27,417)

The mean ELFI score in the INCA3 sample was 19.62 (3.35). Regarding food groups, poultry and vegetables received the highest scores (2.60 (0.58) and 2.29 (0.83), respectively), while dairy foods and added sugars scored the lowest (0.36 (0.81) and 0.67 (0.90), respectively). The total ELFI score showed a positive correlation with ELI (r = 0.44, p < 0.0001), and there was also a positive correlation between the equivalent food groups of both indices, with coefficients ranging from 0.11 to 0.49 (all with p < 0.0001) (Table 3). The correlations between ELFI food groups and the g/d were as expected: positive correlations for the food groups to encourage (coefficients ranging from 0.15 to 0.49, p < 0.0001), while negative correlations for food groups to balance or to limit (coefficients ranging from −0.15 to −0.38, p < 0.0001). The correlation between the ELFI total score and energy was negligent (−0.09) (Figure S1 in Supplementary Material).

**Table 3.**
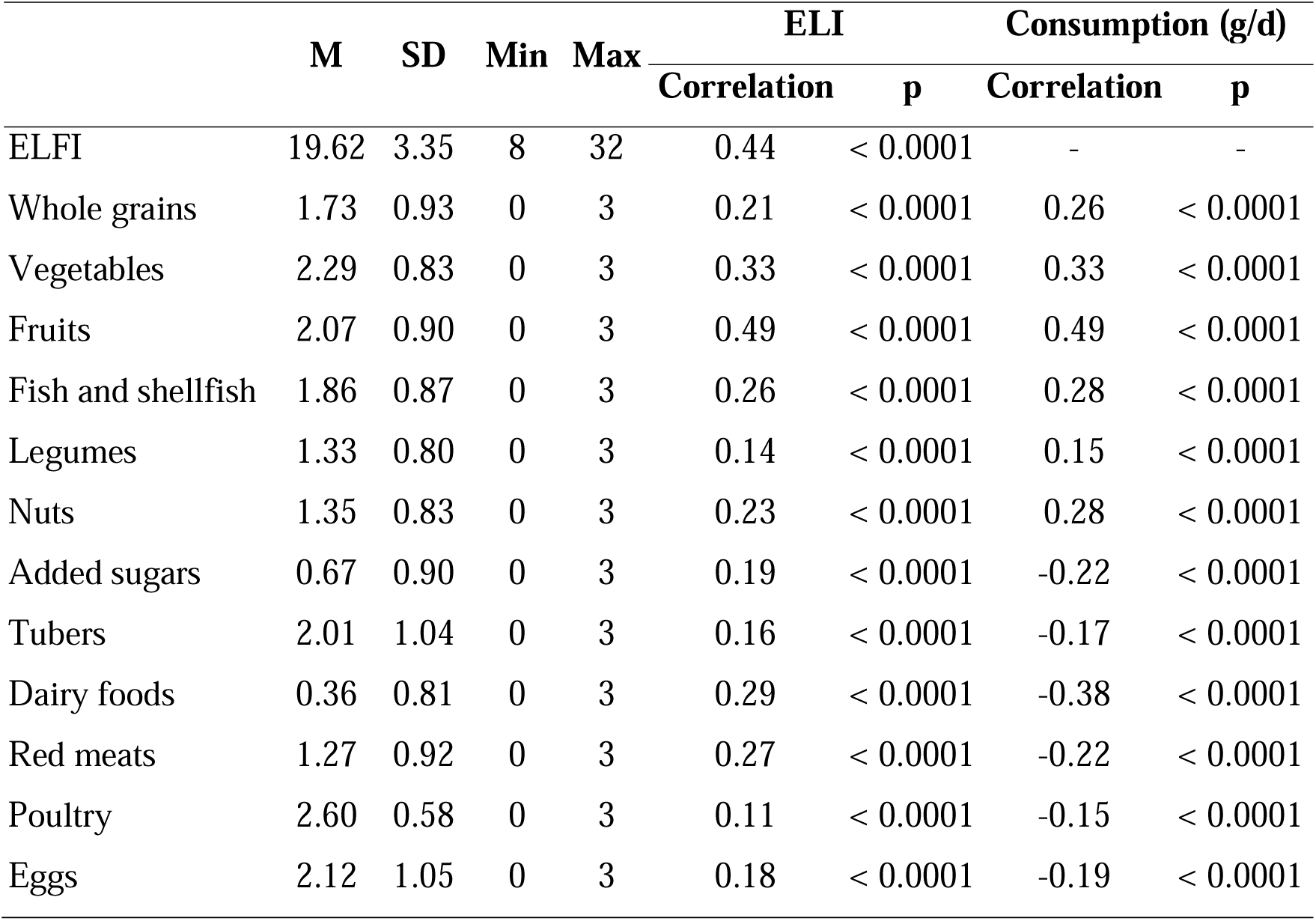
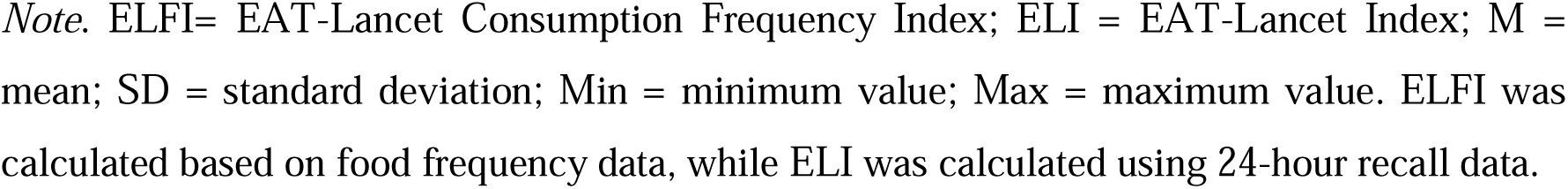
Descriptive statistics of ELFI and correlation with ELI and the habitual food consumption (g/d) from 24-hour recall in the INCA3 survey (n = 1,645)

|  | M | SD | Min | Max | ELI |  | Consumption (g/d) |  |
| --- | --- | --- | --- | --- | --- | --- | --- | --- |
|  |  |  |  |  | Correlation | p | Correlation | p |
| ELFI | 19.62 | 3.35 | 8 | 32 | 0.44 | < 0.0001 | - | - |
| Whole grains | 1.73 | 0.93 | 0 | 3 | 0.21 | < 0.0001 | 0.26 | < 0.0001 |
| Vegetables | 2.29 | 0.83 | 0 | 3 | 0.33 | < 0.0001 | 0.33 | < 0.0001 |
| Fruits | 2.07 | 0.90 | 0 | 3 | 0.49 | < 0.0001 | 0.49 | < 0.0001 |
| Fish and shellfish | 1.86 | 0.87 | 0 | 3 | 0.26 | < 0.0001 | 0.28 | < 0.0001 |
| Legumes | 1.33 | 0.80 | 0 | 3 | 0.14 | < 0.0001 | 0.15 | < 0.0001 |
| Nuts | 1.35 | 0.83 | 0 | 3 | 0.23 | < 0.0001 | 0.28 | < 0.0001 |
| Added sugars | 0.67 | 0.90 | 0 | 3 | 0.19 | < 0.0001 | -0.22 | < 0.0001 |
| Tubers | 2.01 | 1.04 | 0 | 3 | 0.16 | < 0.0001 | -0.17 | < 0.0001 |
| Dairy foods | 0.36 | 0.81 | 0 | 3 | 0.29 | < 0.0001 | -0.38 | < 0.0001 |
| Red meats | 1.27 | 0.92 | 0 | 3 | 0.27 | < 0.0001 | -0.22 | < 0.0001 |
| Poultry | 2.60 | 0.58 | 0 | 3 | 0.11 | < 0.0001 | -0.15 | < 0.0001 |
| Eggs | 2.12 | 1.05 | 0 | 3 | 0.18 | < 0.0001 | -0.19 | < 0.0001 |

*Note.* ELFI= EAT-Lancet Consumption Frequency Index; ELI = EAT-Lancet Index; M = mean; SD = standard deviation; Min = minimum value; Max = maximum value. ELFI was calculated based on food frequency data, while ELI was calculated using 24-hour recall data.

Table 4 presents a comparison of food consumption across ELFI quintiles. Significant positive trends were observed for all food groups categorized as “to encourage”, with the most notable differences seen in the consumption of fruits, vegetables, and nuts. In contrast, significant negative trends were identified for food groups categorized as “to balance and to limit”, particularly concerning added sugars and red meats. Additionally, no variation in poultry consumption was observed from Q1 to Q5.

**Table 4.** Food consumption (g/d) across ELFI quintiles in INCA3 survey (n = 1,645)

| Food groups | Q1 (n = 364) |  | Q5 (n = 385) |  | ANOVA |  | Trend |  |
| --- | --- | --- | --- | --- | --- | --- | --- | --- |
|  | M | SD | M | SD | F | P | Direction | p-trend |
| Whole grains | 11.46 | 23.69 | 23.30 | 40.89 | 11.54 | < 0.0001 | + | < 0.0001 |
| Vegetables | 199.20 | 176.52 | 312.46 | 199.53 | 19.49 | < 0.0001 | + | < 0.0001 |
| Fruits | 116.84 | 128.90 | 248.56 | 169.88 | 50.03 | < 0.0001 | + | < 0.0001 |
| Fish and shellfish | 25.99 | 38.91 | 41.72 | 45.19 | 7.71 | < 0.0001 | + | < 0.0001 |
| Legumes | 12.88 | 36.58 | 8.34 | 20.50 | 4.25 | 0.0020 | + | 0.0017 |
| Nuts | 2.88 | 7.02 | 7.04 | 13.50 | 12.81 | < 0.0001 | + | < 0.0001 |
| Oils | 7.31 | 9.41 | 10.04 | 10.62 | 4.21 | 0.0022 | + | < 0.0001 |
| Fats | 32.33 | 23.19 | 23.62 | 17.89 | 9.67 | < 0.0001 | - | < 0.0001 |
| Added sugars | 62.68 | 45.22 | 41.43 | 38.08 | 15.86 | < 0.0001 | - | < 0.0001 |
| Tubers | 73.09 | 85.93 | 53.10 | 63.62 | 4.45 | 0.0014 | - | < 0.0001 |
| Dairy foods | 236.69 | 185.96 | 202.96 | 147.48 | 2.08 | 0.0812 | - | 0.0026 |
| Red meats | 108.64 | 71.73 | 77.39 | 58.97 | 11.05 | < 0.0001 | - | < 0.0001 |
| Poultry | 30.66 | 37.55 | 30.71 | 36.56 | 2.03 | 0.0884 | +/- | 0.6938 |
| Eggs | 28.11 | 29.06 | 25.56 | 27.81 | 3.51 | 0.0073 | - | 0.0004 |

*Note.* ELFI= EAT-Lancet Consumption Frequency Index; M = mean; SD = standard deviation; SD = standard deviation; ANOVA = Analysis of variance. ELFI was calculated based on food frequency data, while ELI was calculated using 24-hour recall data.

Figure 4 shows the results of the concurrent-criterion validity analysis. Women scored significantly higher on ELFI (β = 0.09, p < 0.0001), as did individuals aged 45-64 and 65-79 compared to those aged 18-44 (β = 0.21, p < 0.0001 and β = 0.19, p < 0.0001, respectively). Higher educational level was also positively associated with ELFI, with individuals in the highest education level group scoring higher than those in the lowest one (β = 0.16, p < 0.0001). Also, those with high-school to two years of higher education scored marginally higher on ELFI (β = 0.06, p = 0.057). Similarly, participants with incomes between €900-€1,850 scored higher than those earning less than €900 (β = 0.10, p = 0.002; β = 0.07, p = 0.042; and β = 0.16, p < 0.0001, respectively). Household size was also related to ELFI: those living alone scored higher than individuals in households of three (β = −0.08, p = 0.010) or more than five members (β = −-0.12, p < 0.0001). Sedentary behavior, regardless of physical activity level, was associated with lower ELFI compared to active and non-sedentary individuals (β = −0.15, p = 0.001 and β = −0.13, p = 0.005, respectively). Smoking was also associated with lower ELFI (β = −0.06, p = 0.019). Finally, although individuals with normal weight tended to score higher in ELFI, this association was not statistically significant.

**Figure 4.**
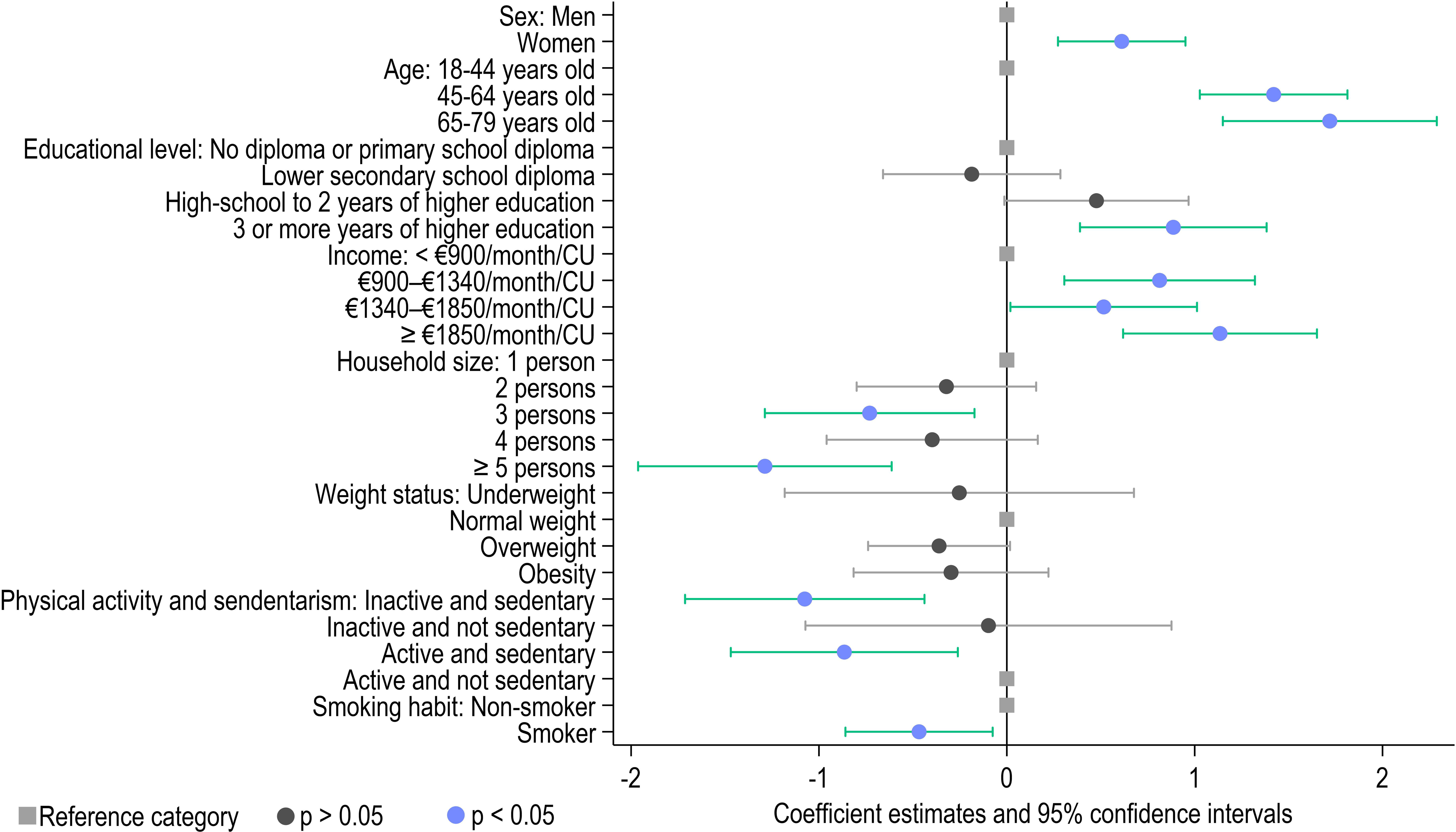
Multivariate regression coefficient plot with confidence intervals illustrating the association of EAT-Lancet Consumption Frequency Index (ELFI) with sociodemographic variables in INCA3 survey (n = 1,645).

When analyzing the bivariate correlations of the ELFI index with nutritional and environmental indicators (Figure 5), ELFI positively correlated with the adequacy of 16 nutrients, including fatty acids, fiber, vitamins (thiamine, folate, vitamins C, D, and E), minerals (magnesium, copper, and manganese), carbohydrates, saturated fatty acids, and sugars. In contrast, three nutrients showed negative correlations with ELFI: zinc, calcium, and niacin. Regarding diet quality, ELFI exhibited positive correlations with all evaluated indicators, except for the DII, which had a negative correlation, indicating that a higher ELFI is associated with a lower inflammatory potential of the diet. ELFI was associated with a reduced environmental impact, showing negative correlations with most environmental pressure indicators, except for water use, which had a positive correlation, and photochemical ozone formation, which was not significant.

**Figure 5.**
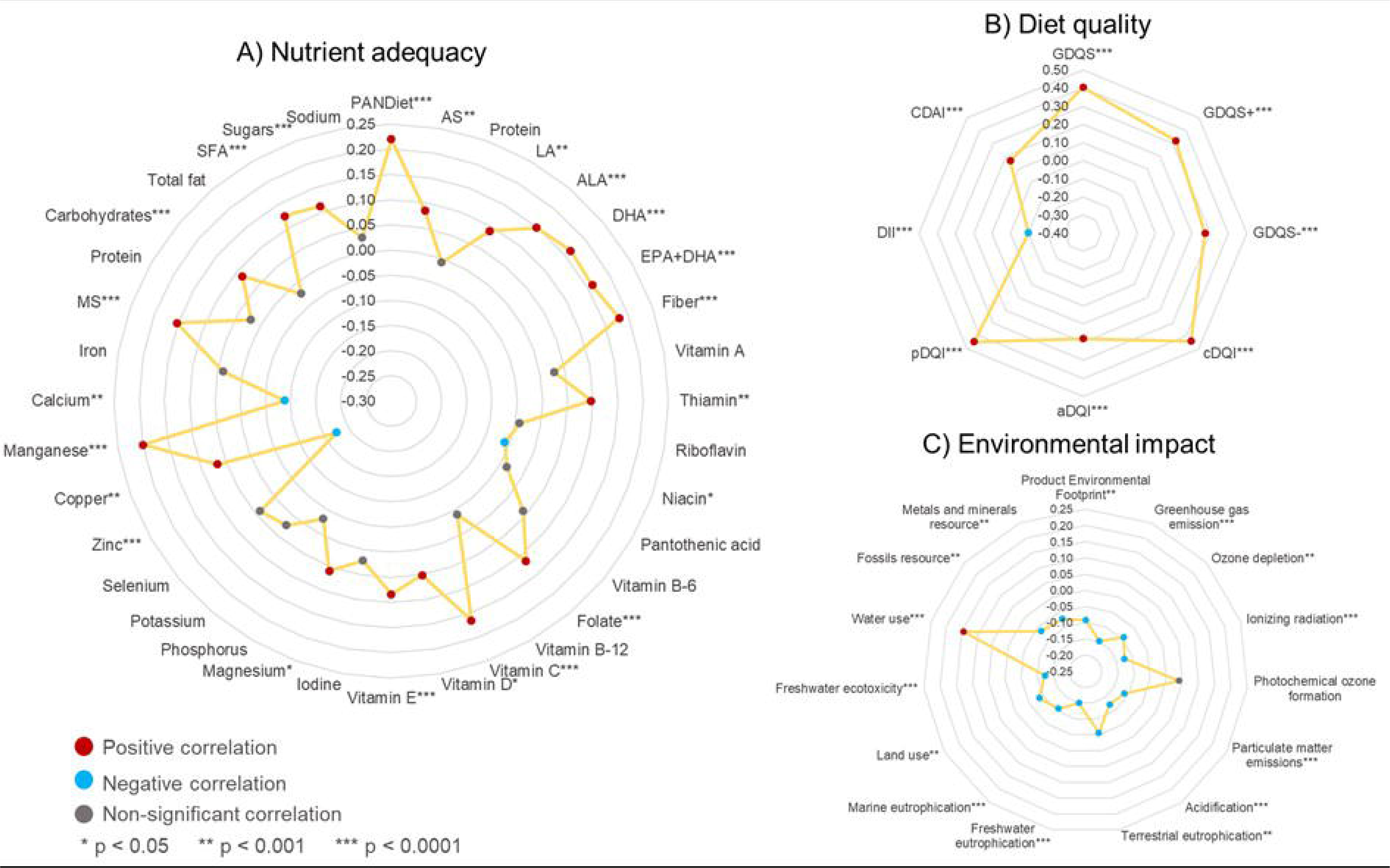
EAT-Lancet Consumption Frequency Index (ELFI) correlations with nutrient adequacy (A), diet quality (B), and environmental impact (C) in INCA3 survey (n = 1,645). Values expressed as Spearman’s correlation coefficients. CDAI = Composite Dietary Antioxidant Index; DQI = Comprehensive Diet Quality Index (comprehensive, animal, or plant); DII = Dietary Inflammatory Index; GDQS = Global Diet Quality Score; PANDiet = Probability of Adequate Nutrient intake Diet score; AS = adequacy subscore; ALA = alpha-linolenic acid; DHA = docosahexaenoic acid; EPA = eicosapentaenoic acid; LA = linoleic acid; MS = moderation subscore; SFA = saturated fatty acids.

Figure 6 displays the results of SEM analysis on nomological validity. The model demonstrated a good fit and was able to explain 90% of data variability. All measured variables showed significant loadings on their respective latent variables (p < 0.0001). Regarding the structural model (i.e., the relationships between the latent variables), expected associations were found, confirming the nomological validity of ELFI. Specifically, “foods to encourage” factor was positively associated with “nutritional health” (β = 0.62, p < 0.0001) and negatively associated with “environmental impact” (β = −0.16, p < 0.0001). Similarly, “foods to balance and limit” factor was positively correlated with “nutritional health” (β = 0.23, p < 0.0001) and negatively correlated with “environmental impact” (β = −0.36, p < 0.0001). Additionally, there was a negative correlation between the “foods to encourage” and “foods to balance and limit” (β = −0.50, p < 0.0001).

**Figure 6.**
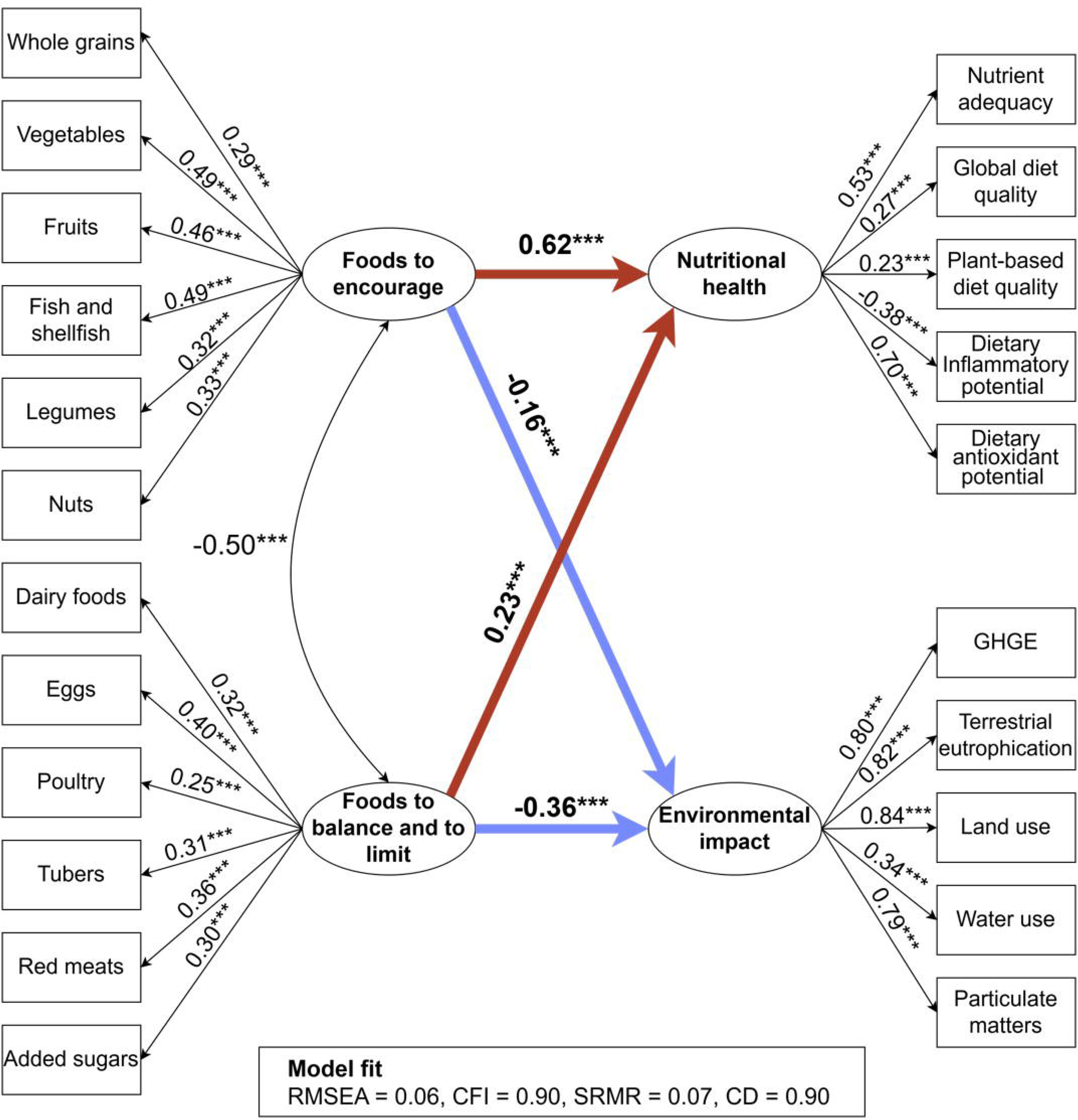
Assessment of nomological validity of the EAT-Lancet Consumption Frequency Index (ELFI): Structural equation model with standardized coefficients in INCA3 survey (n = 1,645). The rectangles represent the observed variables; the elliptic circles symbolize the latent variables. Red arrows indicate positive associations, while blue arrows represent negative associations. For the sake of clarity, errors are not shown. RMSEA = root mean square error of approximation; CFI = comparative fit index; SRMR = standardized root mean square residual; CD = coefficient of determination. ***p < 0.0001.

## 4. Discussion

This study aimed to develop and validate the ELFI, a brief and practical index based on FPQ for assessing relative adherence to the planetary health diet in large-scale multi-country surveys. Our findings demonstrate that the ELFI is both reliable and valid for measuring relative adherence to the planetary health diet proposed by the EAT-Lancet Commission in such surveys. The index showed adequate internal consistency and a robust multidimensional structure, allowing it to capture a broad range of dietary variability. Moreover, ELFI was found to correlate with the ELI and food group consumption from 24-hour dietary recalls, and was associated with indicators of better nutritional health and a lower environmental impact. These results support the applicability of ELFI in large-scale studies. Consequently, our findings offer valuable insights for researchers seeking valid instruments to assess adherence to sustainable and healthy diets in large-scale research settings. To the best of our knowledge, this is the first study to develop and validate an approach based on FPQs that represent the EAT-Lancet food groups.

The average ELFI score in both samples were notably lower than the maximum possible scores, suggesting that few individuals closely follow the general dietary recommendations of the EAT-Lancet planetary health diet. This finding aligns with the results of other indices developed to measure absolute adherence to the planetary health diet in terms of quantity levels. For example, the ELI index had an average score of 18 (out of a maximum of 42) (28), the WISH index had a mean of 46 (out of 130) (52), and the PHDI had a mean of 60 (out of 150) (10). Moreover, the scores by food groups in this study align with dietary trends observed in Europe and France, lending support to the validity of the ELFI index (53, 54).

The reliability and structural validity of the ELFI index was corroborated using a large-scale multi-country survey conducted in 27 European countries. The high internal consistency, evidenced by an α coefficient exceeding 0.80, suggests that ELFI’s food groups share common covariance and collectively measure a cohesive underlying concept. Each food group contributes uniquely to the overall measure, with no gain in reliability after removal and no redundancy (25). The internal consistency of ELFI, both at the global and subscore levels, observed in this study aligns with that of other indices assessing healthy and sustainable diets (55, 56). For instance, other dietary indices based on the EAT-Lancet have reported α above 0.50 (10, 57, 58). Although a minimum α of 0.70 is generally recommended, nutritional metrics often require more flexible thresholds (above 0.50) due to the complexity and multidimensionality of dietary constructs (10).

The structural validity of ELFI was confirmed, revealing a bidimensional structure: one factor for “foods to encourage” and another factor for “foods to balance and to limit”. Testing unidimensional, bidimensional, and bifactor models supported both the validity of the two subscores and the overall total score. These models confirmed that “foods to encourage” and “foods to balance and to limit” provide distinct, independent information while enhancing ELFI’s global measure. Thus, the total score offers a general measure, while each subscore yields specific insights into dietary behaviors with implications for health and environmental impact. Multidimensionality has also been demonstrated in other indices based on the EAT-Lancet planetary health diet, including the PHDI, which consists of several components (10, 58). Additionally, ELFI’s bidimensional structure aligns with other indices, such as the Healthful Eating Indicator, which includes “recommended foods” and “discretionary foods” subscores to assess dietary behaviors in at-risk groups (59). ELFI’s structure also aligns with the GDQS, which comprises GDQS+ for 16 healthy food groups and GDQS-for nine unhealthy food groups (37). Recent studies suggest that GDQS+ and GDQS-scores associate with positive health outcomes, such as reduced risks of metabolic syndrome, cardiovascular disease, obesity, and diabetes (60–62). Thus, future studies should further explore associations between ELFI and health outcomes.

Convergent validity was established based on the agreement of the ELFI with valid criteria derived from 24-hour dietary recall data. In this regard, ELFI showed a moderate positive correlation with ELI, along with positive correlations between the food groups of both indices. ELI was selected to test the convergent validity of the ELFI due to its methodological alignment and robust validation in relation to health outcomes. ELI was developed using dietary data obtained from food diaries and FFQ as part of the Malmö Diet and Cancer study, which involved a large cohort of 22,421 Swedish adults aged 45 to 73 (28). Thus, this large sample and the dietary assessment framework provide a reliable basis for comparison with ELFI. Moreover, ELI’s structure covering 14 food components grouped into emphasized and restricted categories, with daily quantities scored through a graded system, parallels the conceptual framework of ELFI (12). This similarity enhances the relevance of ELI as a benchmark for measuring absolute adherence to EAT-Lancet planetary health diet. Importantly, ELI has demonstrated associations with positive health outcomes, including reduced all-cause mortality, lower mortality from cancer and cardiovascular disease, and a decreased risk of type 2 diabetes and mental disorders (28, 63–65). Furthermore, adherence to the ELI has been associated with reduced dietary costs among lower socioeconomic groups, largely due to decreased consumption of red meats, dairy foods, and added sugars (66). These correlations with health outcomes strengthen ELI’s reliability and provide a robust reference point for ELFI validation for measuring relative adherence to the planetary health diet. The moderate agreement between ELFI and ELI is expected, as both assess adherence to the same dietary pattern; however, methodological differences may introduce variability. In this sense, ELFI focuses on consumption frequency, whereas ELI considers intake quantity, which can lead to some differences (67). Nevertheless, the correlation of 0.44 indicates a moderate association between the indices, suggesting that frequency and quantity represent complementary dimensions of dietary adherence (68). From a research perspective, ELFI may be more suitable for population-based research, where consumption frequency is a key indicator and a rapid and efficient assessment is required, while ELI is better suited for detailed analyses where precise intake quantification is critical.

Furthermore, ELFI was independent of energy intake, consistent with the findings for other EAT-Lancet indices (6, 10, 12). A positive correlation with energy intake could have biased ELFI scores, favoring high-calorie diets regardless of their nutritional value or environmental sustainability (25). Conversely, a strong negative correlation would also require careful interpretation, as very low-calorie diets may not meet essential nutritional needs. Overall, such correlations could lead to misleading assessments, with diets that are either excessive or insufficient in calories, but not necessarily balanced or sustainable, appearing to have high diet quality scores (25).

On the other hand, ELFI scores varied across sociodemographic groups, demonstrating its concurrent-criterion validity. These results are in accordance with previous research conducted in France (12). For instance, the Programme National Nutrition Santé – guidelines score (PNNS-GS), an index designed to reflect the 2017 French dietary recommendations, showed higher scores in the same sociodemographic groups (69). Likewise, the validation study of the Sustainable Diet Index in France revealed that participants with higher scores were predominantly women and university graduates (70). Other studies have reported that most of the EAT-Lancet indices exhibit significant associations with the analyzed sociodemographic variables (12). In general terms, research has indicated that factors such as being women, being older, having a higher educational level, higher income, a physically active life style, and non-smoking status are linked to higher scores in the ELI (28, 65), Planetary Health Diet Index (PHDI) (71–73), EAT-Lancet Diet Index (ELDI) (74,75), and EAT-Lancet Diet Score (ELDS) (76,77). Although a marginal association was identified between normal weight and higher ELFI scores, this relationship did not achieve statistical significance. This may be attributed to the use of self-reported weight and height for BMI calculations, which can introduce self-report bias. This bias may be especially pronounced in higher BMI categories, where individuals often misreport their weight (78).

A key strength of this study is that it demonstrated the nomological validity of the ELFI index in relation to both nutritional health and environmental impact. Nomological validity examines whether the instrument’s scores correlate with other measures that, theoretically, they should relate to (50). After examining the bivariate correlations of the ELFI index with individual measures, a nomological model was developed, demonstrating that higher scores in “foods to encourage” and “foods to balance and to limit” were associated with improved nutritional health (in terms of nutrient adequacy and diet quality), as well as a reduction in environmental impact. While correlations were significant and aligned in the expected direction, the “foods to encourage” factor had its strongest positive effect on nutritional health, while the “foods to balance and to limit” had a greater effect on reducing environmental impact. This difference in effects between food groups may be explained by the design of the planetary health diet. Foods in the “to encourage” groups are typically nutrient-dense (e.g., fruits, vegetables, legumes, and nuts) sources of essential vitamins, minerals, and fiber, thus directly enhancing dietary quality and nutrient adequacy (79). In contrast, foods in the “to balance and to limit” groups include items with a higher environmental impact, such as red meats and dairy foods, which are more resource-intensive and produce greater emissions (80,81).

These findings should be considered alongside certain limitations. The INCA3 survey, conducted between 2014 and 2015, may not capture recent dietary pattern changes, nonetheless, it remains the most current, representative food consumption dataset for France. Additionally, the INCA3’s FPQ does not include vegetable oils and animal fats, limiting the complete validity assessment; however, these food groups were assessed and validated in the FEAST survey. The cross-sectional design of both INCA3 and FEAST further restricts the estimation of predictive validity and associations with health outcomes. However, a major strength of this study is the large sample size used to develop ELFI, incorporating data from 27 European countries, which enhances the reliability of the findings.

Limitations of the Agribalyse v.3.1.1 database should also be acknowledged, including its lack of data on soil carbon storage, biodiversity, organic versus non-organic food distinctions, and incomplete information on water use and waste indicators (82,83). Some limitations are inherent to the use of the FPQ and the scoring system applied in this study. Notably, the consumption frequency reported by participants may not fully reflect their habitual food intake, a limitation commonly observed in studies of this type, potentially introducing bias in the assessment of actual consumption (20). Regarding quantile-scoring system, cutoffs developed are population-specific, limiting the generalization (84). Also, it assumes equal spacing between consumption frequency levels, which may overlook the real variability within each level and limit the detection of more detailed or extreme consumption patterns.

Despite this, our findings indicate that relative adherence, as measured by ELFI, correlates with absolute adherence, as measured by ELI. This suggests that both measures capture similar aspects of dietary behavior, making ELFI a potentially valuable approach for assessing relative adherence to planetary health diet in large-scale studies, particularly when obtaining absolute adherence data is challenging.

Furthermore, as with other indices based on the EAT-Lancet diet, ELFI tends to assign higher scores to certain special diets, such as veganism and vegetarianism, which could compromise the balanced intake of some nutrients (85). To address this limitation, we avoided restrictive scoring applying a more flexible criteria to the food groups to be balanced or limited, with the exception of added sugars and animal fats, where specific restrictions were maintained. It is important to highlight that zinc, calcium, and niacin adequacy showed an inverse association with ELFI in the bivariate analysis. This is noteworthy given that nutrient-rich foods such as red meats and dairy foods were categorized as “to encourage”. Although these associations were weak (i.e., coefficients <0.20), these findings align with existing discussions and criticisms of the one-size-fits-all approach of the EAT-Lancet planetary health diet (12, 86). This dietary model has been debated for its potential to contribute to nutrient deficiencies, particularly in populations with diverse dietary patterns and specific physiological needs (86). Research assessing the micronutrient adequacy of the EAT-Lancet diet has indicated that it may not provide sufficient levels of iron, calcium, and zinc, raising potential public health concerns (85). In consequence, these findings highlight the need for future research to refine this dietary framework, ensuring it adequately supports nutrient intake while balancing health and environmental considerations. This issue is expected to be addressed in the next version of the EAT-Lancet diet, which may contribute to the development of more nuanced adherence scores, such as ELFI (87).

## Conclusion

To our knowledge, ELFI is the first dietary index specifically designed to assess relative adherence to the planetary health diet through a FPQ approach. ELFI enables evaluation of individuals’ alignment with the principles of this diet by employing a quantile-based scoring system derived from population consumption data. This method supports meaningful comparisons by categorizing individuals into levels of relative adherence and provides flexibility by adapting to diverse populations and dietary contexts without the need for fixed cut-off points. In terms of validity, ELFI demonstrates significant correlations with ELI, a validated measure of the planetary health diet, and is associated with improved nutritional adequacy, higher dietary quality, and reduced environmental impact. Furthermore, the index exhibited strong reliability and structural validity for large-scale surveys, with a multidimensional design that delivers both an overall score and two sub-scores reflecting specific dietary dimensions. Future research should explore ELFI in diverse contexts and examine its associations with health and environmental outcomes to further validate its applicability. Additionally, future research may benefit from updating ELFI in line with future revisions to the planetary health diet, particularly regarding its nutritional adequacy.

Overall, ELFI presents a robust and easy-to-implement tool for assessing relative adherence to sustainable and healthy diets in large-scale studies, contributing valuable insights into dietary patterns in the context of sustainability and health research.

## Statements

### Author contribution

AM and EV conceptualized and designed the study, and drafted the manuscript. AM conducted the statistical analyses. FC, FM, and AMM coordinated the FEAST questionnaire design, led the survey implementation and critically reviewed the manuscript draft. AJ supervised the project and contributed a critical review of the draft. FDC and MM contributed in the methodology for index development and validation. All authors read and approved the final manuscript. AM and EV held primary responsibility for the final content.

### Data Availability

Data from the Third French Individual and National Food Consumption Survey (INCA3) in available on the data.gouv.fr platform at: https://www.data.gouv.fr/fr/datasets/donnees-de-consommations-et-habitudes-alimentaires-de-letude-inca-3/. Data on the environmental impacts of foods consumed in France is available on the agribalyse.ademe.fr platform. FEAST anonymous survey data will be made available coherently with the FEAST Data management Plan: https://feast2030.eu/sites/default/files/feast/resources_files/2024/FEAST_WP1_DMP-PlanDel1.2_D_20240528_V2.pdf.

## Abbreviations

AIC: Akaike Information Criterion
ALA: alpha-linolenic acid
ANCOVA: Analysis of Variance
ANSES: French Agency for Food, Environmental and Occupational Health and Safety
AS: adequacy subscore
BIC: Bayesian Information Criterion
BMI: Body Mass Index
CD: Coefficient of Determination
CDAI: Composite Dietary Antioxidant Index
cDQI: Comprehensive Diet Quality Index
CFA: confirmatory factor analysis
CFI: Comparative Fit Index
CU: Consumption Unit
DHA: docosahexaenoic acid
DII: Dietary Inflammatory Index
EFA: exploratory factor analysis
EPA: eicosapentaenoic acid
ELD-I: EAT-Lancet Diet Index
ELDS: EAT-Lancet Diet Score
ELFI: EAT-Lancet Consumption Frequency Index
ELI: EAT-Lancet dietary index
FEAST: Food systems that support transitions to hEalthy And Sustainable dieTs project
FFQ: food frequency questionnaires
FPQ: Food propensity questionnaires
GDQS: Global Diet Quality Score
INCA3: French Third Individual and National Study on Food Consumption Survey
KMO: Kaiser-Meyer-Olkin
LA: linoleic acid
LCA: Life Cycle Assessment
METs: metabolic equivalent of task
MS: moderation subscore
PANDiet: Probability of Adequate Nutrient intake Diet score
PHDI: Planetary Health Diet Index
RMSEA: Root Mean Square Error of Approximation
SEM: structural equation modeling
SFA: saturated fatty acids
SRMR: Standardized Root Mean Square Residual.

