## Supplementary materials for "Assessing sustainable and healthy diets in large-scale surveys: validity and applicability of a dietary index based on a brief food group propensity questionnaire representing the EAT-Lancet planetary health diet"

**EAT-Lancet Index**

The EAT-Lancet Index (ELI) was developed using data from 22,421 Swedish adults aged 45 to 73 who participated in the Malmö Diet and Cancer cohort between 1991 and 1996, which included food diaries and food frequency questionnaires. The ELI covers 14 food components categorized as emphasized and restricted foods, with daily gram quantities assessed through a semi-quantitative scoring system: from 0 to 3 points. This index has been related with reduced all-cause mortality, decreased mortality from cancer and cardiovascular diseases. Furthermore, it was observed that higher adherence to this index is associated with a lower risk of type 2 diabetes in Swedish adults, and this relationship is dose-response and independent of genetic susceptibility. The following table presents the food components of the ELI, proving examples of food items and a more detailed explanation of the scoring.

| **Food components** | **Food items** | **Scoring** | | | |
| --- | --- | --- | --- | --- | --- |
|  |  | **3 points** | **2 points** | **1 point** | **0 points** |
| **Vegetables** | All vegetables except legumes. | >300 | 200–300 | 100–200 | <100 |
| **Fruits** | Fruits and berries. | >200 | 100–200 | 50–100 | <50 |
| **Unsaturated oils** | All plant oils and plant margarines. | >40 | 20–40 | 10–20 | <10 |
| **Legumes** | Dry beans, lentils, peas, soy. Peas, lentils, beans, tofu, soy containing meat replacement products. | >75 | 37.5–75 | 18.75–37.5 | <18.75 |
| **Nuts** | Peanuts or tree nuts. All nuts and seeds including peanuts, nut mixes such as almond paste. | >50 | 25–50 | 12.5–25 | <12.5 |
| **Whole grains** | Whole grains (e.g., cereals, rolled oats, crispbread) and whole grain foods (e.g., pastas, doughs and breads). | >232 | 116–232 | 58–116 | <58 |
| **Fish** | Fatty fish, lean fish, fish products, shellfish. | >28 | 14–28 | 7–14 | <7 |
| **Beef and lamb** | Beef, lamb, minced meat with pork and lamb, processed meats with beef and lamb including sausages. | <7 | 7–14 | 14–28 | >28 |
| **Pork** | Pork, minced meat of pork, processed meats with pork including ham, bacon, and sausages. | <7 | 7–14 | 14–28 | >28 |
| **Poultry** | Chicken, turkey, duck, goose, and other poultry. | <29 | 29–58 | 58–116 | >116 |
| **Eggs** | Boiled eggs, fried eggs and eggs in dishes such as omelet and pie. | <13 | 13–25 | 25–50 | >50 |
| **Dairy** | Whole milk or derivative equivalents. Regular milk, low-fat milk, yoghurt and other fermented milk products, hard cheese, soft cheese, cream, butter, butter-based spreads. All dairy foods were expressed as of milk equivalents. Equivalency factor: whole milk 1.0, Cheese 5.0, cream 2.7 and butter 6.5. | <250 | 250–500 | 500–1000 | >1000 |
| **Potatoes** | Boiled potatoes, fried potatoes, deep fried potatoes, potatoes included in dishes such as potato salad. | <50 | 50–100 | 100–200 | >200 |
| **Added sugar** | Sucrose and monosaccharides except sugars in fruits and vegetable. | <31 | 31–62 | 62–124 | >124 |

Stubbendorff A, Sonestedt E, Ramne S, Drake I, Hallström E, Ericson U. Development of an EAT-Lancet index and its relation to mortality in a Swedish population. Am J Clin Nutr. 2022;115(3):705-716.

**PANDiet scoring system**

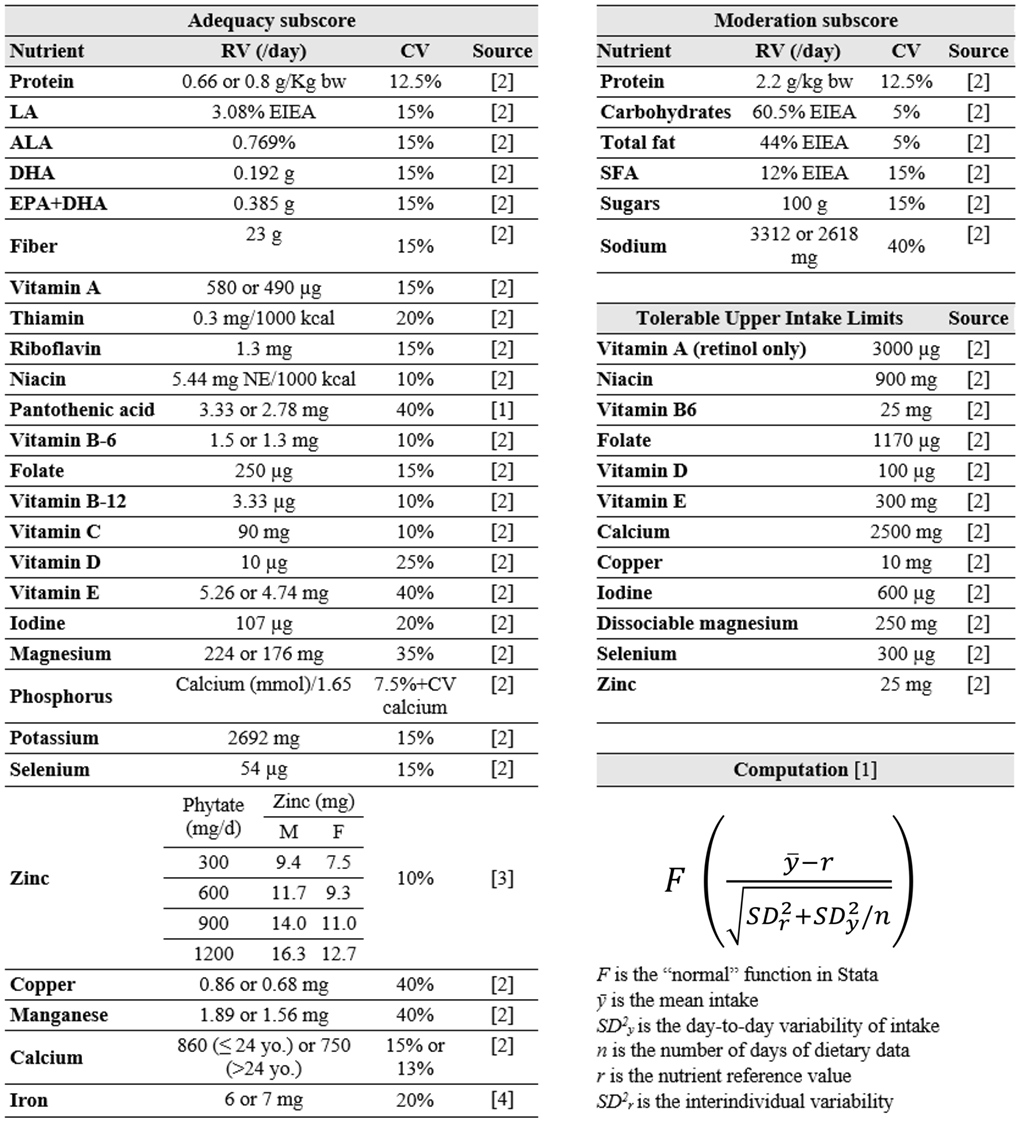
The PANDiet is a measure of individual diet quality expressed as a 100-point score that determines the probability that overall nutrient intake is adequate [1]. The PANDiet is calculated as the mean of two sub-scores: The Adequacy Subscore (i.e., the probability of that intake to be adequate), and the Moderation Subscore (i.e., the probability of not exceeding intake of nutrients that should be limited). The total PANDiet score ranges from 0 to 100, with a higher score indicating a better nutritional quality of the diet. The following table presents the PANDiet scoring system.

*Note*. DHA and EPA+DHA are weighted by a factor of 1/2 as DHA is present twice. Niacin equivalents are calculated as the sum of dietary niacin and 1/60 dietary tryptophan. ALA, alpha-linolenic acid; bw, body weight; CV, coefficient of variation; DHA, docosahexaenoic acid; EIEA, energy intake excluding alcohol; EPA, eicosapentaenoic acid; LA, linoleic acid; NE, niacin equivalent; SFA, saturated fatty acids.

[1] Verger EO, Mariotti F, Holmes BA, Paineau D, Huneau JF. Evaluation of a Diet Quality Index Based on the Probability of Adequate Nutrient Intake (PANDiet) Using National French and US Dietary Surveys. PLoS ONE. 2012;7(8).

[2] Salomé M, Mariotti F, Nicaud MC, Dussiot A, Kesse-Guyot E, Maillard MN, Huneau JF, Fouillet H. Eropean Journal of Nutrition. 2022;61(4):1991-2002.

[3] EFSA (2014). Draft Scientific Opinion on Dietary Reference Values for Zinc. EFSA Journal 2014. Available online: https://www.efsa.europa.eu/sites/default/files/consultation/140514%2C0.pdf. Accessed 30 Nov 2023.

[4] ANSES (2021). Les références nutritionnelles en vitamines et minéraux. In: Anses—Agence nationale de sécurité sanitaire de l’alimentation, de l’environnement et du travail. Available online: https://www.anses.fr/fr/content/les-r%C3%A9f%C3%A9rences-nutritionnelles-en-vitamines-et-min%C3%A9raux. Accessed 30 Nov 2023.

**Global Diet Quality Score**

The Global Diet Quality Score (GDQS) is a diet quality assessment tool focusing on 25 food groups considered critical for nutrient intake and/or risk of non-communicable diseases. The score is composed of 16 food groups categorized as healthy (scored positively to encourage higher intake), seven groups classified as unhealthy (scored negatively to penalize higher levels of intake), and two food groups that are considered unhealthy only if consumed in excessive amounts (score 0 if intake is low or excessive). The total score ranges from 0 to 49. Also, two additional GDQS-related sub-metrics are calculated: the GDQS+ (only for food groups considered healthy), which ranges from 0 to 32; and the GDQS- (only for food groups considered unhealthy or harmful in excessive amounts), which ranges from 0 to 17. The following table presents the GDQS scoring system.

| **Food group** | **Food items** | | | **Categories of consumption** | | | | | | | **Point values** | | | | | | |
| --- | --- | --- | --- | --- | --- | --- | --- | --- | --- | --- | --- | --- | --- | --- | --- | --- | --- |
|  |  |  |  | **1** | **2** | | **3** | | **4** | | **1** | | **2** | | **3** | | **4** |
| ***GDQS+*** | | | |  |  | |  | |  | |  | |  | |  | |  |
| ***Healthy*** |  | | |  |  | |  | |  | |  | |  | |  | |  |
| Citrus fruits | Fruits in the genus Citrus | | | <24 | 24–69 | | >69 | |  | | 0 | | 1 | | 2 | |  |
| Deep orange fruits | Fruits containing ≥20 retinol equivalents/100 g | | | <25 | 25–123 | | >123 | |  | | 0 | | 1 | | 2 | |  |
| Other fruits | Fruits not belonging in the other fruit categories | | | <27 | 27–107 | | >107 | |  | | 0 | | 1 | | 2 | |  |
| Dark green leafy vegetables | Leafy vegetables containing >120 retinol equivalents/100 g | | | <13 | 13–37 | | >37 | |  | | 0 | | 2 | | 4 | |  |
| Cruciferous vegetables | Vegetables in the family Brassicaceae | | | <13 | 13–36 | | >36 | |  | | 0 | | 0.25 | | 0.5 | |  |
| Deep orange vegetables | Non-tuberous vegetables containing ≥120 retinol equivalents/100 g | | | <9 | 9–45 | | >45 | |  | | 0 | | 0.25 | | 0.5 | |  |
| Other vegetables | Vegetables not belonging in the other vegetable categories | | | <23 | 23–114 | | >114 | |  | | 0 | | 0.25 | | 0.5 | |  |
| Legumes | Legumes and foods derived from legumes. | | | <9 | 9–42 | | >42 | |  | | 0 | | 2 | | 4 | |  |
| Deep orange tubers | Tuberous vegetables containing ≥120 retinol equivalents/100 g | | | <12 | 12–63 | | >63 | |  | | 0 | | 0.25 | | 0.5 | |  |
| Nuts and seeds | Nuts, seeds, and products derived | | | <7 | 7–13 | | >13 | |  | | 0 | | 2 | | 4 | |  |
| Whole grains | Whole grains and whole-grain products. | | | <8 | 8–13 | | >13 | |  | | 0 | | 1 | | 2 | |  |
| Liquid oils | All types of oils that are liquid at room temperature. | | | <2 | 2–7.5 | | >7.5 | |  | | 0 | | 1 | | 2 | |  |
| Fish and shellfish | Fish (processed or unprocessed) and seafood (e.g., shellfish). | | | <14 | 14–71 | | >71 | |  | | 0 | | 1 | | 2 | |  |
| Poultry and game meat | Unprocessed poultry and game. | | | <16 | 16–44 | | >44 | |  | | 0 | | 1 | | 2 | |  |
| Low fat dairy | Reduced or naturally low-fat dairy products (≤2% milk fat). | | | <33 | 33–132 | | >132 | |  | | 0 | | 1 | | 2 | |  |
| Eggs | All types of eggs. Does not include mayonnaise | | | <6 | 6–32 | | >32 | |  | | 0 | | 1 | | 2 | |  |
| **Food group** | **Food items** | **Categories of consumption** | | | | | | | | **Point values** | | | | | | | |
|  |  | **1** | **2** | | | **3** | | **4** | | **1** | | **2** | | **3** | | **4** | |
| ***GDQS-*** | |  |  | | |  | |  | |  | |  | |  | |  | |
| ***Unhealthy in excessive amounts*** | |  |  | | |  | |  | |  | |  | |  | |  | |
| High fat dairy | High fat milk and dairy products (>2% milk fat), except butter, ice cream and whipped cream. | <35 | 35–142 | | | >142–734 | | >734 | | 0 | | 1 | | 2 | | 0 | |
| Red meat | Unprocessed red meat belonging to domesticated animals (i.e., not game), including organs. | <9 | 9–46 | | | >46 | |  | | 0 | | 1 | | 0 | |  | |
| ***Unhealthy*** |  |  |  | | |  | |  | |  | |  | |  | |  | |
| Processed meat | Processed red meat, poultry, or game, including organs. | <9 | 9–30 | | | >30 | |  | | 2 | | 1 | | 0 | |  | |
| Refined grains and baked goods | Refined grains and refined grain products. | <7 | 7–33 | | | >33 | |  | | 2 | | 1 | | 0 | |  | |
| Sweets and ice cream | Sugar-sweetened foods that are not beverages (e.g., sugar, sweeteners, whipped cream). | <13 | 13–37 | | | >37 | |  | | 2 | | 1 | | 0 | |  | |
| Sugar-sweetened beverages | Sweetened drinks that do not contain any fruit juice (e.g., sodas, energy drinks, sports drinks). | <57 | 57–180 | | | >180 | |  | | 2 | | 1 | | 0 | |  | |
| Juice | Unsweetened or sweetened drinks that are at least partly composed of fruit juice. | <36 | 36–144 | | | >144 | |  | | 2 | | 1 | | 0 | |  | |
| White roots and tubers | Tuberous vegetables with <120 retinol equivalents/100 g. | <27 | 27–107 | | | >107 | |  | | 2 | | 1 | | 0 | |  | |
| Purchased deep fried foods | Deep fried foods fried in an amount of fat or oil sufficient to cover the food. | <9 | 9–45 | | | >45 | |  | | 2 | | 1 | | 0 | |  | |

Bromage S, Batis C, Bhupathiraju SN, Fawzi WW, Fung TT, Li Y, Deitchler M, Angulo E, Birk N, Castellanos-Gutiérrez A, He Y, Fang Y, Matsuzaki M, Zhang Y, Moursi M, Gicevic S, Holmes MD, Isanaka S, Kinra S, Sachs SE, Stampfer MJ, Stern D, Willett WC. Development and Validation of a Novel Food-Based Global Diet Quality Score (GDQS). J Nutr. 2021;151(12 Suppl 2):75S-92S.

**Comprehensive Diet Quality Index**

The Comprehensive Diet Quality Index (cDQI) is a recently developed measure that ranges from 0 to 85 and is obtained by adding two sub-indexes: the plant-based diet quality index (pDQI) and the animal-based diet quality index (aDQI). The main purpose of this metric is to distinguish between the consumption of plant and animal products that are considered healthy and those that are not. The cDQI aims to provide a comprehensive assessment of diet quality, considering both plant and animal components. The two sub-indices are calculated using thresholds based on literature evidence or crude consumption quintiles. The following table presents the cDQI scoring system.

|  |  | **Max score (5)** | **Min Score (0)** |
| --- | --- | --- | --- |
| **Plant-based Diet Quality Index (pDQI)** | | | |
| ***Healthful*** | |  |  |
|  | Wholegrain products | ≥ 45g per 1000/kcal | No consumption |
|  | Vegetables (excluding potatoes) | ≥ 125g per 1000/kcal | No consumption |
|  | Fruit | ≥ 125g per 1000/kcal | No consumption |
|  | Nuts, seeds, legumes | ≥ 14,175g per 1000/kcal | No consumption |
|  | Vegetable oils | ≥ 14.86g per 1000/kcal | < 4.13g per 1000/kcal |
|  | Coffee, tea | ≥477.35g per 1000/kcal | < 91.72g per 1000/kcal |
| ***Unhealthy*** | |  |  |
|  | Fruit juices | No consumption | ≥79.38g per 1000/kcal |
|  | Refined grains | <54g per 1000/kcal | ≥129g per 1000/kcal |
|  | Potatoes | No consumption | ≥ 35g per 1000/kcal |
|  | Sugar-sweetened beverages | No consumption | ≥ 226.8g per 1000/kcal |
|  | Sweets and desserts | < 14.98g per 1000/kcal | ≥ 40.32g per 1000/kcal |
| Range pDQI | | **0 to 55** |  |
| **Animal-based Diet Quality Index (aDQI)** | | | |
| ***Healthful*** | |  |  |
|  | Fish, seafood | ≥ 14,175g per 1000/kcal | No consumption |
|  | Dairy | ≥ 312g per 1000/kcal | No consumption |
|  | Poultry | ≥ 17.09g per 1000/kcal | < 3.12g per 1000/kcal |
| ***Unhealthy*** | |  |  |
|  | Processed meat | No consumption | ≥ 28,35g per 1000/kcal |
|  | Red meat | No consumption | ≥ 45,36g per 1000/kcal |
|  | Eggs | < 1.78g per 1000/kcal | ≥ 8.62g per 1000/kcal |
| Range aDQI | | **0 to 30** |  |
| **cDQI Total Range** | | **0 to 85** |  |

Brunin J, Allès B, Péneau S, Reuzé A, Pointereau P, Touvier M, et al. Do individual sustainable food purchase motives translate into an individual shift towards a more sustainable diet? A longitudinal analysis in the NutriNet-Santé cohort. Clean Responsib Consum. (2022) 5:100062.

**Dietary Inflammatory Index**

The Dietary Inflammatory Index (DII) is a population-based index derived from the literature for the purpose of measuring the overall impact of diet on inflammatory potential. It is based on the individual inflammatory effects of up to 45 dietary parameters. Unlike single compound or single food approaches, a strategic advantage of the DII is that it allows the study of the dietary matrix and the complex interactions of nutrients and compounds present in foods, as well as overall dietary patterns.

The specific steps for the calculation were as follows:

1) dietary intake data were compared with the global standard dietary intake database, and a Z-score was calculated for each nutrient based on the mean and standard deviation of that nutrient intake:

$$Z score= \frac{individual reported intake-global daily mean intake}{global standard deviation}$$

2) the Z-scores were converted into centered proportions,
3) the centered proportion for each dietary parameter was multiplied by the specific inflammatory effect score to obtain a DII score,
4) DII scores were summed for all dietary parameters to obtain the total DII score.

Since its release, the DII index has undergone rigorous validations in both cross-sectional and longitudinal studies. These validations have yielded consistent results indicating that higher DII scores are linked to proinflammatory dietary patterns. In addition, a significant association has been observed between elevated DII scores and elevated plasma concentrations of proinflammatory cytokines over time and health outcomes.

The following table presents the DII scoring system for the 34 parameters used in the current study.

| **Food parameter** | **Overall inflammatory effect score** | **Global daily mean intake (units/d)** | **SD** |
| --- | --- | --- | --- |
| Alcohol (g) | −0.278 | 13.98 | 3.72 |
| Vitamin B12 (μg) | 0.106 | 5.15 | 2.70 |
| Vitamin B6 (mg) | −0.365 | 1.47 | 0.74 |
| β-Carotene (μg) | −0.584 | 3718 | 1720 |
| Carbohydrate (g) | 0.097 | 272.2 | 40.0 |
| Energy (kcal) | 0.180 | 2056 | 338 |
| Total fat (g) | 0.298 | 71.4 | 19.4 |
| Fiber (g) | −0.663 | 18.8 | 4.9 |
| Folic acid (μg) | −0.190 | 273.0 | 70.7 |
| Garlic (g) | −0.412 | 4.35 | 2.90 |
| Ginger (g) | −0.453 | 59.0 | 63.2 |
| Fe (mg) | 0.032 | 13.35 | 3.71 |
| Mg (mg) | −0.484 | 310.1 | 139.4 |
| MUFA (g) | −0.009 | 27.0 | 6.1 |
| Niacin (mg) | −0.246 | 25.90 | 11.77 |
| n-3 Fatty acids (g) | −0.436 | 1.06 | 1.06 |
| n-6 Fatty acids (g) | −0.159 | 10.80 | 7.50 |
| Onion (g) | −0.301 | 35.9 | 18.4 |
| Protein (g) | 0.021 | 79.4 | 13.9 |
| PUFA (g) | −0.337 | 13.88 | 3.76 |
| Riboflavin (mg) | −0.068 | 1.70 | 0.79 |
| Saffron (g) | −0.140 | 0.37 | 1.78 |
| Saturated fat (g) | 0.373 | 28.6 | 8.0 |
| Se (μg) | −0.191 | 67.0 | 25.1 |
| Thiamin (mg) | −0.098 | 1.70 | 0.66 |
| Turmeric (mg) | −0.785 | 533.6 | 754.3 |
| Vitamin A (RE) | −0.401 | 983.9 | 518.6 |
| Vitamin C (mg) | −0.424 | 118.2 | 43.46 |
| Vitamin D (μg) | −0.446 | 6.26 | 2.21 |
| Vitamin E (mg) | −0.419 | 8.73 | 1.49 |
| Zn (mg) | −0.313 | 9.84 | 2.19 |
| Pepper (g) | −0.131 | 10.00 | 7.07 |
| Thyme/oregano (mg) | −0.102 | 0.33 | 0.99 |
| Rosemary (mg) | −0.013 | 1.00 | 15.00 |

Shivappa N, Steck SE, Hurley TG, Hussey JR, Hébert JR. Designing and developing a literature-derived, population-based dietary inflammatory index. Public Health Nutr. 2014;17(8):1689-96.

**Composite Dietary Antioxidant Index**

The Composite Dietary Antioxidant Index (CDAI) is a comprehensive measure that assesses an individual's antioxidant profile by combining several dietary antioxidants, such as vitamins A, C and E, manganese, selenium and zinc. Its development was based on the cumulative effect of anti-inflammatory indicators related to various health problems mediated by oxidative processes.

Widely used to evaluate the antioxidant capacity of dietary components, the CDAI is calculated from the levels of vitamins A, C and E, as well as manganese, selenium and zinc. The formula used to calculate the CDAI has been previously validated and is expressed as follows:

$$CDAI=\sum_{i=1}^{n=6} \frac{Total nutrient intake-mean}{SD}$$

Here, the total nutrient intake reflects the daily intake of antioxidant for each individual; the mean represents the mean of the antioxidant in the whole sample, while the standard deviation (SD) indicates the variability of the antioxidant in the whole sample.

Wang L, Yi Z. Association of the Composite dietary antioxidant index with all-cause and cardiovascular mortality: A prospective cohort study. Front Cardiovasc Med. 2022;9:993930.

**Correlation between ELFI and total daily energy intake**

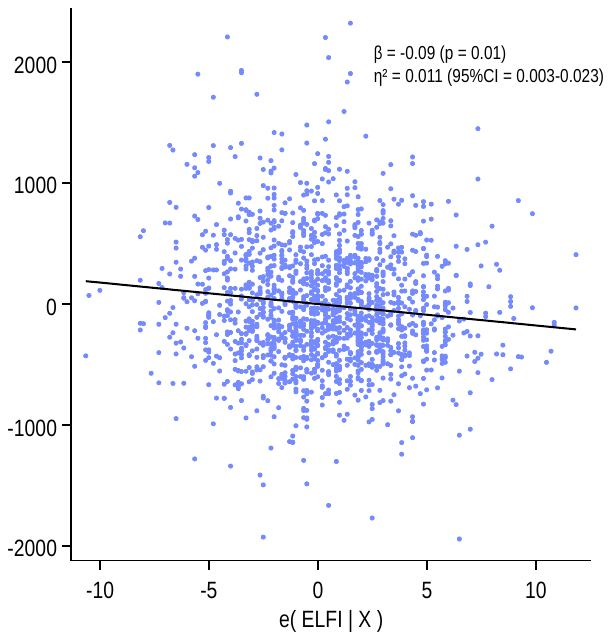

**Figure S1**. Added variable plot demonstrating the correlation between ELFI and daily energy intake adjusted by sex and age.
